# Altered Gastrointestinal Interoception in Anorexia Nervosa Predicts Relapse

**DOI:** 10.1101/2025.09.16.25335842

**Authors:** Charles Verdonk, Keller Mink, Emily Choquette, Scott E. Moseman, Ahmad Mayeli, Jennifer L. Stewart, Martin P. Paulus, Ryan Smith, Sahib S. Khalsa

**Affiliations:** Laureate Institute for Brain Research, Tulsa, Oklahoma, United States; UMR VIFASOM, Université de Paris, Paris, France; French Armed Forces Biomedical Research Institute, Brétigny-sur-Orge, France; Oxley College of Health Sciences, University of Tulsa, Tulsa, Oklahoma, United States; Laureate Eating Disorders Program, Tulsa, Oklahoma, United States; University of Pittsburgh, Pittsburgh, Pennsylvania, United States; Department of Psychiatry and Biobehavioral Sciences, Semel Institute for Neuroscience and Human Behavior, David Geffen School of Medicine, University of California, Los Angeles, California, United States

## Abstract

Anorexia nervosa (AN) is a deadly psychiatric disorder, yet relapse remains common after weight restoration and no objective tools exist to track recovery. AN involves profound disturbances of gastrointestinal interoception, but no clinical tests assess this dysfunction. In this single-blind, randomized-crossover study, 62 weight-restored females with restrictive AN and 57 matched healthy comparisons completed a gastrointestinal detection task using an ingestible vibrating capsule while behavioral, electroencephalographic, and peripheral physiological signals were recorded. AN participants showed reduced perceptual accuracy and higher miss-rates despite intact neural and physiological responses. Computational modeling revealed stronger prior beliefs against perceiving gut sensations, diminished interoceptive precision, and maladaptive learning. Capsule stimulation also induced larger hunger increases in AN. Initial priors, response bias, and stomach unpleasantness predicted six-month relapse, while miss-rate and precision shifts predicted symptom severity. These findings reveal mechanistic disruptions in gastrointestinal interoception that predict relapse, offering scalable biomarkers to personalize treatment and prevent recurrence.

## INTRODUCTION

Anorexia nervosa (AN) is a severe and often enduring psychiatric disorder with one of the highest mortality rates, with suicide as a leading cause of death^1,2^. Despite treatment, relapse rates are alarmingly high—up to 50% within one year of weight restoration^3,4^. This persistent risk underscores the critical need for an improved understanding of the pathophysiological mechanisms underlying the disorder and for objective biomarkers that can track how patients respond to treatment and predict their long-term outcomes.

Interoception, the nervous system’s process of sensing and interpreting signals from within the body^5^, has been consistently implicated in the symptomatology of AN^6,7^ and is often regarded as a candidate mechanism underlying AN^8,9^. Empirical work has implicated abnormal interoception, particularly in the cardiac and respiratory domains, as a contributor to symptoms such as appetite dysregulation, anxiety, and distorted body image^10–12^. However, much less is known about interoception of gastrointestinal (GI) signals in AN despite its central role in satiety, meal anticipation, and visceral discomfort—features intimately tied to AN symptomatology^13^. Methodological limitations have historically hampered GI-focused research, with most available tools being either invasive (e.g., insertion of inflatable balloons or electrical probes into the digestive tract) or poorly suited for repeated use in clinical populations (e.g., water loading tests; see^14^ for discussion).

To address this methodological gap, we previously validated a minimally invasive approach involving an ingestible vibrating capsule delivering controlled mechanosensory stimulation to the stomach and adjacent segments of the gut^15^. In healthy individuals, vibrating capsules reliably evoked non-aversive gut sensations and elicited a neural signature measurable via electroencephalography (EEG), including parieto-occipital event-related potentials (ERPs) that correlated with perceptual accuracy^15^. We further incorporated a Bayesian computational model of interoceptive inference to estimate latent perceptual processes such as prior beliefs, sensory precision, and learning from internal signals, demonstrating associations between interoceptive precision, learning rates, and both parieto-occipital and frontal ERPs^16^.

Computational accounts of AN have suggested that symptoms such as exaggerated fullness or visceral discomfort may result from aberrant interoceptive inference, specifically, the over-weighting of prior expectations and/or under-weighting of sensory input^17–19^. In addition, biases in interoceptive learning may be crucial for maintaining these maladaptive expectations. Such mechanisms could help explain the persistence of disordered eating behaviors even after weight normalization, and may reflect a broader rigidity in updating beliefs based on changing internal states^20^.

In the current study, we tested whether inpatient individuals with weight-restored AN exhibit disrupted GI interoception across behavioral, neural, and computational domains. We predicted that compared to healthy comparisons (HCs), individuals with AN would show reduced perceptual accuracy and diminished ERP responses to gut stimulation. We further hypothesized that individuals with AN would display stronger prior beliefs against detecting gut sensations, lower interoceptive precision, and impaired learning from mechanosensory input from the gut. Finally, we tested whether these interoceptive parameters would predict relapse and severity of symptoms six months after discharge, thereby identifying potential biomarkers of prognosis and treatment response.

## METHODS

### 1. Participants

Participants included weight-restored females with restrictive AN and sex- and age-matched HCs, recruited from the Laureate Eating Disorder Program and the surrounding community, respectively.

Inclusion criteria for the AN group were: 1) female sex, 2) age between 13 and 40 years, 3) Body mass index (BMI) ≥ 18.5 (to minimize starvation-related physiological confounds ^21,22^), and 4) primary diagnosis of restrictive AN as confirmed by the treating psychiatrist. Comorbid generalized anxiety disorder, specific phobia, dysthymia or major depressive disorder were permitted, due to their high prevalence in AN^23^. Participants taking psychiatric medications were allowed into the study, provided the dose was stable for > 1 week (Table S1 in Supplementary Methods 1. Participants). Exclusion criteria included active suicidality, a history of severe purging, pregnancy or lactation, or serious GI illness (Table S2 in Supplementary Methods 1. Participants).

HCs were recruited from the local community and screened using the MINI clinical interview^24^ to rule out DSM-5 diagnosis. The study was approved by the Western Institutional Review Board. All participants provided written informed consent before participation and were financially compensated for their involvement. ClinicalTrials.gov identifier: #NCT05111977.

### 2. Experimental session

The experimental protocol consisted of a 30-minute resting baseline, followed by two blocks of vibratory stimulations delivered by a swallowed ingestible capsule (Vibrant Ltd) (Figure 1). Approximately 3 minutes post-ingestion, participants performed the GI detection task while seated. Stimulation blocks (normal and enhanced) were counterbalanced and consisted of approximately 60 vibratory stimulations of 3 seconds each^15^. Participants were instructed to press and hold a button upon detecting capsule-induced sensations and to release the button once the sensation ceased (Supplementary Methods 2. Experimental protocol). A digital stethoscope affixed to the abdomen verified vibration timing^15^ (Supplementary Methods 2. Experimental protocol).

**Figure 1.**
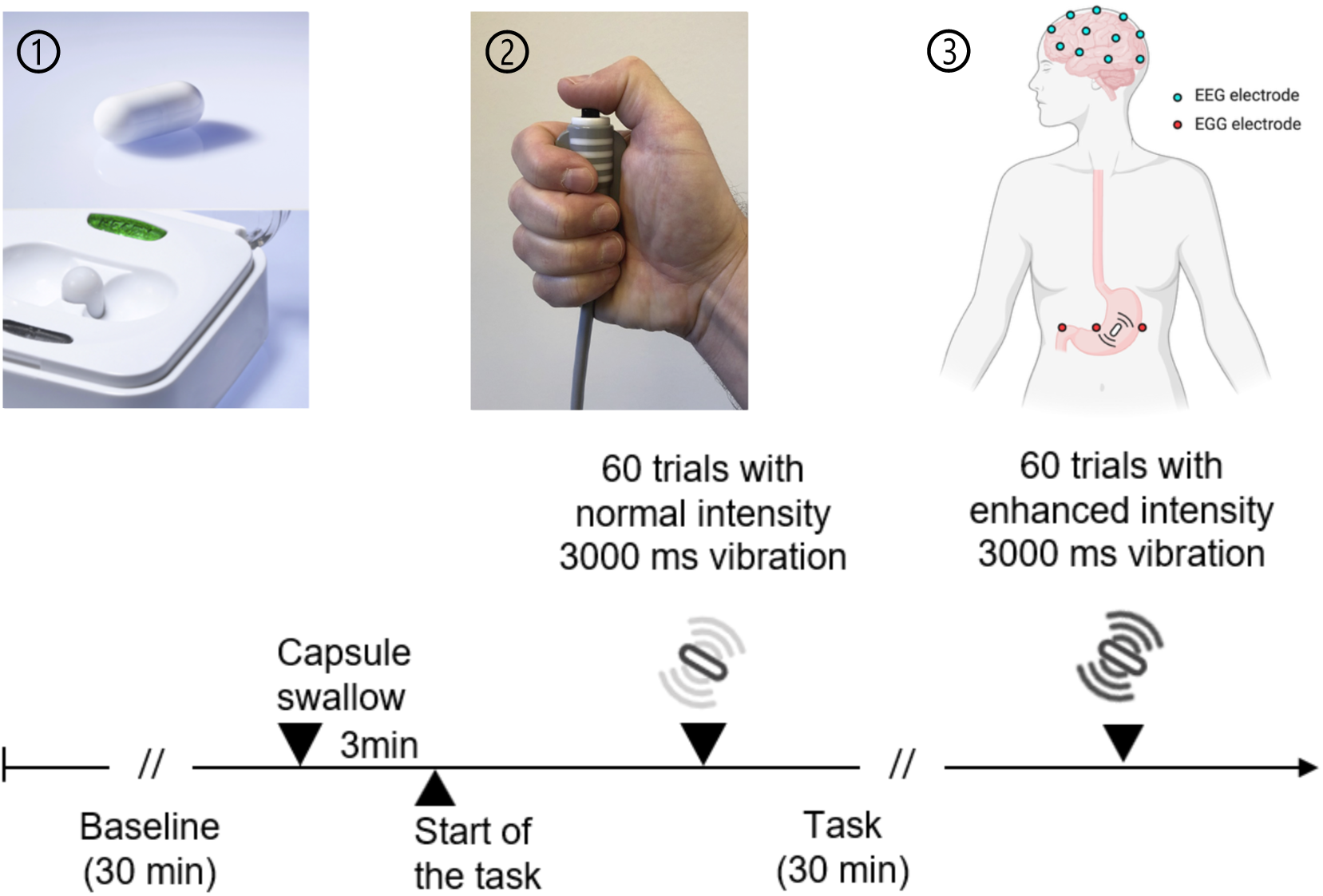
Experimental setup. The gastrointestinal (GI) detection task included two counterbalanced blocks of stimulation (normal and enhanced) generated by the vibrating capsule (Vibrant Ltd). The capsule was activated just before swallowing by placing it in the activation base (①). Each block included approximately 60 stimulations, each lasting 3 seconds. Participants were instructed to continuously attend to their stomach/digestive system and to press and hold a button (②) as soon as they felt a sensation attributed to the capsule and to release the button as soon as the sensation ended. During the GI detection task, neural (electroencephalogram), gastric (electrogastrogram), cardiac (electrocardiogram), galvanic (skin conductance), and subjective responses were recorded simultaneously (③). Abbreviations: ms, milliseconds; min, minutes.

### 3. Follow-up

AN participants were followed remotely at 1-, 3-, and 6-months post-experiment. This report focused on 6-month relapse and symptom outcomes as this window provided the greatest sensitivity for detecting full relapse following treatment using standardized definitions^3,4^.

### 4. Measures

#### 4.1. Behavioral measures and self-report measures

Perceptual accuracy was quantified via normalized A-prime (Supplementary Methods 3. Behavioral measures and self-report measures), a non-parametric signal detection index analogous to 𝑑′, appropriate for small-trial paradigms^25,26^. Response bias (range: −1 to +1) captured general button press tendencies (Supplementary Methods 3. Behavioral measures and self-report measures). Additional metrics included the miss rate (not included in the normalized A-prime formula) and response time (latency from vibration onset to button press). Before and after the task, participants completed visual analog scale ratings assessing intensity and valence of stomach/digestive, breath, and heartbeat sensations; muscle tension (intensity only); and current hunger, thirst, and urges to urinate/defecate (Supplementary

Methods 3. Behavioral measures and self-report measures).

#### 4.2. Electroencephalographic measures

Scalp electroencephalographic (EEG) data were collected in a noise isolated room using a 32-channel cap (Brain Products GmBH, Germany) arranged according to the international 10-20 system, with 31 channels for EEG and one for electrocardiogram (ECG) recording. EEG data were sampled at 1000 Hz.

Preprocessing was completed using EEGLAB (toolbox version 19.0^27^), and custom scripts in Matlab 2021a (Mathworks^®^). Data were downsampled (250 Hz), notch filtered at 60 Hz, and bandpass filtered (0.1 Hz to 80 Hz) using an eight-order Butterworth band-rejection filter, and subsequently cleaned via Independent Component Analysis (ICA) to remove eyeblinks and saccadic eye movements, muscle activity, and motion-related artifacts. Independent components (ICs) identified as artifact-related based on their spectral/topographic features were removed using the decision criteria of the aE-REMCOR approach^28^ and the rtICA approach^29^. On average, one IC was removed per participant (standard deviation: 2; range: 0–19). EEG data were then referenced to the average of the mastoid electrodes (TP9 and TP10), epoched from the 200 milliseconds (ms) prior to the 3000 ms post-onset of each vibration, and baseline corrected using the pre-stimulus mean (i.e., 200 ms before the vibration onset) to address signal drifts commonly observed in EEG recordings^30^. In addition to onset-locked analyses, we also epoched the EEG data relative to the vibration offset, using the same baseline correction and epoch window (i.e., −200 to +3000 ms from offset).

#### 4.3. Computational modeling

Participant responses were fit using a validated Bayesian model of interoceptive inference developed previously for this task^16^. Across the different parameterizations of this model that were tested and compared, parameters estimated for each participant included some or all of the following: (a) Interoceptive precision (𝐼𝑃), reflecting the expected precision or reliability of the afferent interoceptive signal; (b) Difference in 𝐼𝑃 (𝐼𝑃_*diff*_) across blocks with differing vibration intensity (normal vs. enhanced); (c) Initial prior belief about the likelihood (i.e., frequency) of vibrations (𝑝𝑉); and (d) Learning rates for vibration (𝜂_$_) and non-vibration (𝜂_%$_) trials, reflecting the degree to which expectations change based on whether vibrations are (or are not) presented within a given timeframe. Parameters were estimated using a Variational Bayes optimization algorithm^31,32^. A detailed description of the model is available in the Supplement (Supplementary Methods 4. Computational modeling).

#### 4.4. Peripheral physiological measures

Recordings included baseline, normal-, and enhanced-intensity vibration stimulation periods.

##### 4.4.1. Electrogastrogram data

In line with our previous work^15^, we calculated the absolute power for four gastric frequency ranges: normogastria [2.5–3.5 cycles per minute (cpm)], tachygastria [3.75–9.75 cpm], bradygastria [0.5–2.25 cpm], and total power [0.5–11 cpm] (Supplementary Methods 5.

Processing of peripheral physiological data).

##### 4.4.2. Cardiac data

Consistent with our prior research^15^, we computed the following indices: (1) The standard deviation of R–R intervals (SDNN), a widely used measure of overall heart rate (HR) variability, which has been proposed to reflect autonomic regulation of HR^33^; (2) The phasic-HR, which captures transient heart rate changes induced by capsule vibration. For each period, it was calculated as the difference in HR between the 3-second (s) vibratory stimulation period and the 3-s pre-stimulation period (i.e., the 3 seconds immediately preceding vibration onset). To facilitate baseline comparisons, 60 pseudo-vibration onsets (each of 3-second duration) were introduced into the 30-minute baseline recording, with the first 2 minutes of the baseline excluded to ensure a physiological steady state; and (3) The tonic-HR, also referred to as resting HR, was calculated by averaging HR over 60-s windows (Supplementary Methods 5. Processing of peripheral physiological data).

##### 4.4.3. Skin conductance data

Phasic changes in skin conductance responses (SCRs) to capsule stimulation were analyzed using Continuous Deconvolution Analysis^34^. Following our previous work^15^, we quantified SCRs as the logarithmic transformation of the maximum phasic activity during the 3-second vibration period, relative to the 3-second pre-stimulus period, applying a 0.01 micro Siemen (µS) threshold^35,36^. To assess SCR variations relative to resting physiological conditions, we used the same pseudo-events from the baseline period, as in the cardiac data analysis (Supplementary Methods 5. Processing of peripheral physiological data).

#### 4.5. Definition of 6-month AN illness status

AN illness outcomes at 6-months were categorized into relapse, remission, or partial recovery based on based on self-reported symptoms, behaviors, and BMI, following prior operational criteria^4^ (Supplementary Methods 6. Illness status definitions).

### 5. Statistical analyses

#### 5.1. Cross-sectional

##### 5.1.1. Behavioral, self-report, computational, and peripheral physiological data

Linear mixed effects (LME) models examined group (AN vs. HC), block (normal vs. enhanced), age, and BMI as fixed effects and participant as a random effect. Models were applied to behavioral, physiological, and computational measures, with age and BMI included as covariates. These statistical analyses were performed using R (version 4.2.1^37^)

(Supplementary Methods 7. Statistical analysis).

##### 5.1.2. Electroencephalogram analysis

Non-parametric cluster-based permutation tests assessed ERP amplitude differences across groups and conditions^38,39^. This data-driven method allowed us to compare groups across all electrodes at each time point, effectively addressing the issue of multiple comparisons (Supplementary Methods 7. Statistical analysis). Group differences (AN versus HC) were analyzed separately within each block (normal and enhanced). Cluster permutation was conducted within the whole vibration time window (−200 ms to +3000 ms) using custom scripts in Matlab 2021a and the Fieldtrip toolbox (version 20171022^40^).

Average ERP amplitude (avERP) was calculated across midline parieto-occipital electrodes (P3, P4, O1, O2, Pz, Oz, CP1, CP2, and POz) and across time points (364 to 740 ms following vibration onset). This spatiotemporal window was identified in the present study as the common EEG marker of gut mechanosensation across diagnostic groups (AN and HCs; see Figures 3A-B), and is consistent with the spatiotemporal electrode findings reported in our previous work^15^. We then explored the relationship between neural, behavioral, computational, physiological, and subjective outcomes to examine multilevel associations of interoceptive processing across diagnostic groups using Spearman correlation analyses.

**Figure 2.**
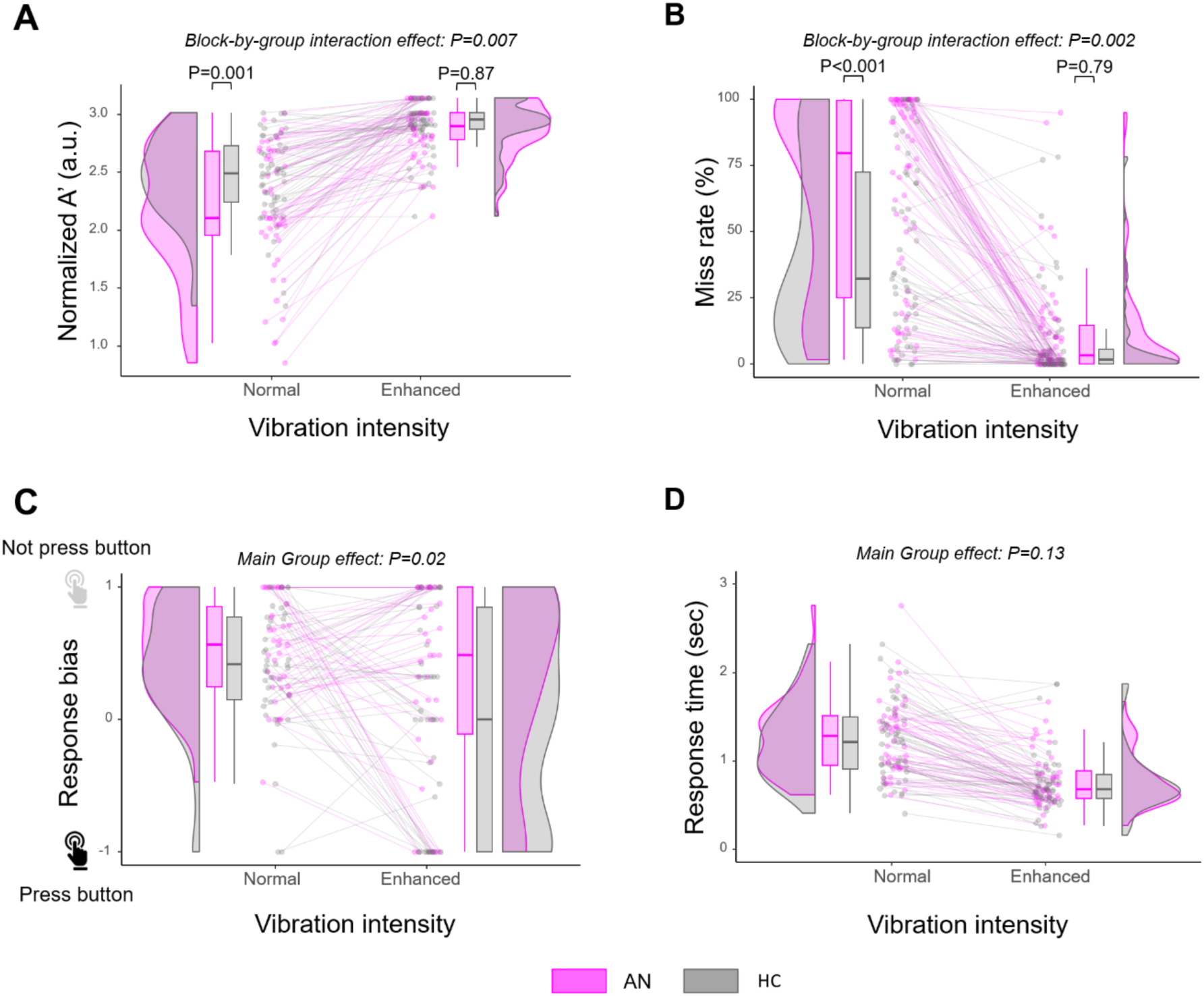
Reduced interoceptive sensitivity to vibratory gut stimulation in AN. AN participants exhibited significantly lower interoceptive accuracy, as measured by normalized A-prime **(A)**, and higher miss rates **(B)** in detecting normal but not enhanced stimulations compared with HCs. **(C)** A significant group effect was found for response bias, with AN participants being less likely to press the button than HCs across blocks. **(D)** Response times did not differ significantly between groups. The plots display the distribution of individual data points, corresponding boxplots, and individual participant values. Abbreviations: AN, anorexia nervosa; HC, healthy comparison; sec - seconds.

**Figure 3.**
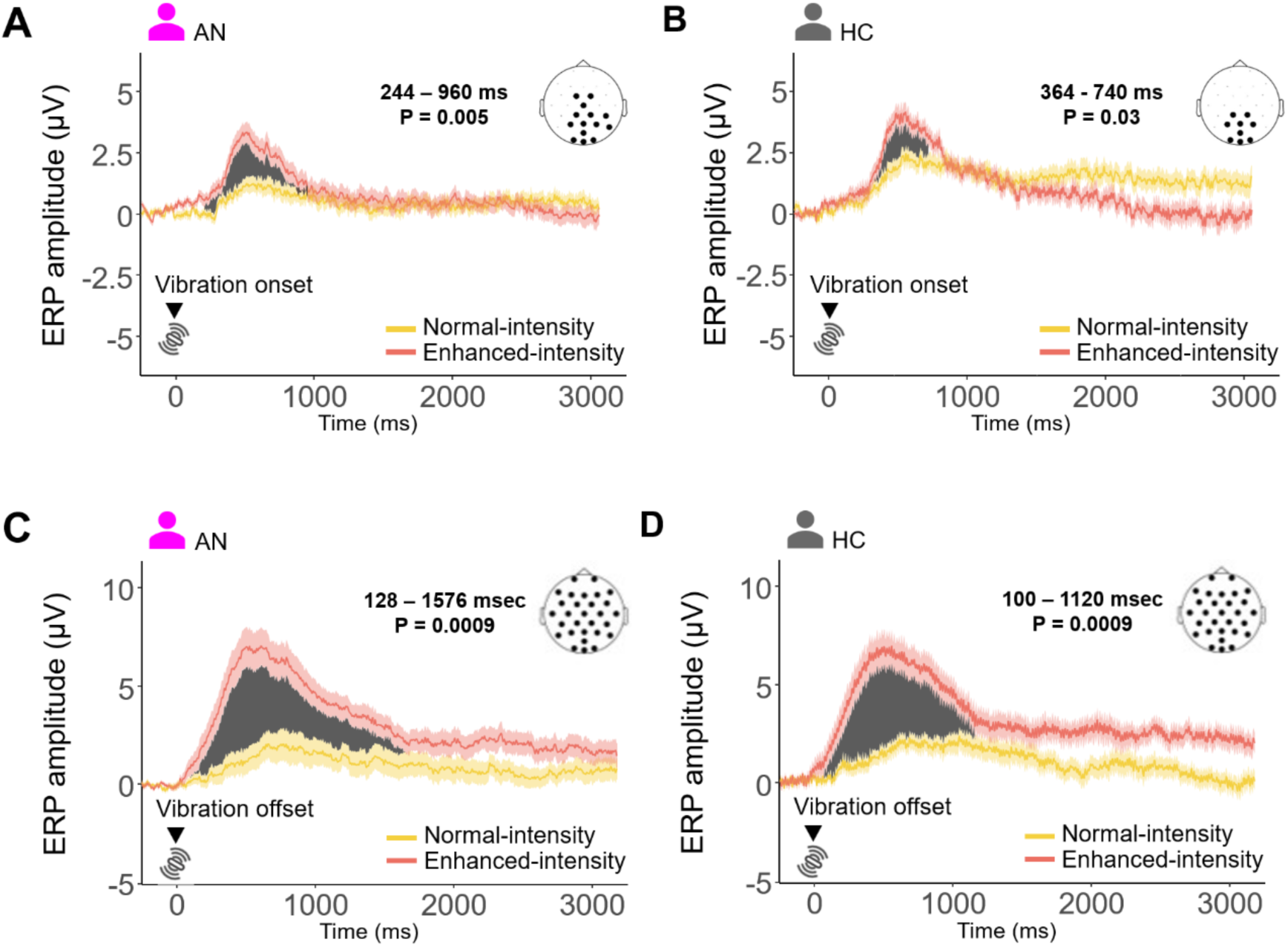
Increased onset and offset event-related potential (ERP) amplitudes in AN and HC during vibratory mechanosensory gut stimulation. Enhanced-intensity stimulations elicited significantly larger ERP amplitudes compared to normal-intensity stimulations in both groups. **(A)** In the AN group, onset-locked ERP amplitudes were significantly greater during enhanced-intensity stimulation (Cohen’s d=0.54), with effects observed in centro-right parieto-occipital electrodes. **(B)** In HCs, onset-locked ERP amplitudes were also significantly greater during enhanced-intensity stimulation (Cohen’s d=0.53), with a partially overlapping set of parieto-occipital electrodes. **(C)** In the AN group, offset-locked ERP amplitudes were significantly larger during enhanced stimulation across all electrodes. **(D)** In HCs, offset-locked ERP amplitudes were likewise significantly larger during enhanced stimulation, with effects spanning all electrodes. The gray shaded area highlights the temporal window where a significant difference was observed between normal- and enhanced-intensity vibrations. Error bars indicate standard error of the mean. Abbreviations: AN, anorexia nervosa; HC, healthy comparison.

### 5.2. Longitudinal analysis

In the AN group, logistic and linear regressions evaluated whether pre-discharge task parameters predicted 6-month relapse outcomes or global scores on the Eating Disorder Examination Questionnaire (EDE-Q), a standard and widely accepted tool for capturing global eating disorder severity^41^. Illness status at 6 months was assessed remotely using operational criteria adapted from our previous standardized definitions^4^, which classified participants as Full relapse, Partial relapse, Partial remission, Full remission, Partial recovery, or Full recovery (Supplementary Methods 6. Illness status definitions). For the logistic models, categories were dichotomized as “Full relapse” vs “Not full relapse” (aggregating Partial relapse, Partial remission, Full remission, Partial recovery, and Full recovery) to address limited variance and ensure stable estimation. Predictors in the logistic regression were standardized by dividing their values by two within-group standard deviations^42^. No scaling was applied in the linear models. Age and BMI were included as covariates in all predictive models.

## RESULTS

For the cross-sectional analysis, 119 female participants with analyzable data were included, comprising 62 individuals with AN restricting subtype and 57 HCs (Table 1; Figure S1 in Supplementary Methods 1. Participants). For the longitudinal analysis, 6-month follow-up data were available for 54 individuals with AN (dropout rate: 13%; Figure S1 in Supplementary Methods 1. Participants).

**Table 1.**
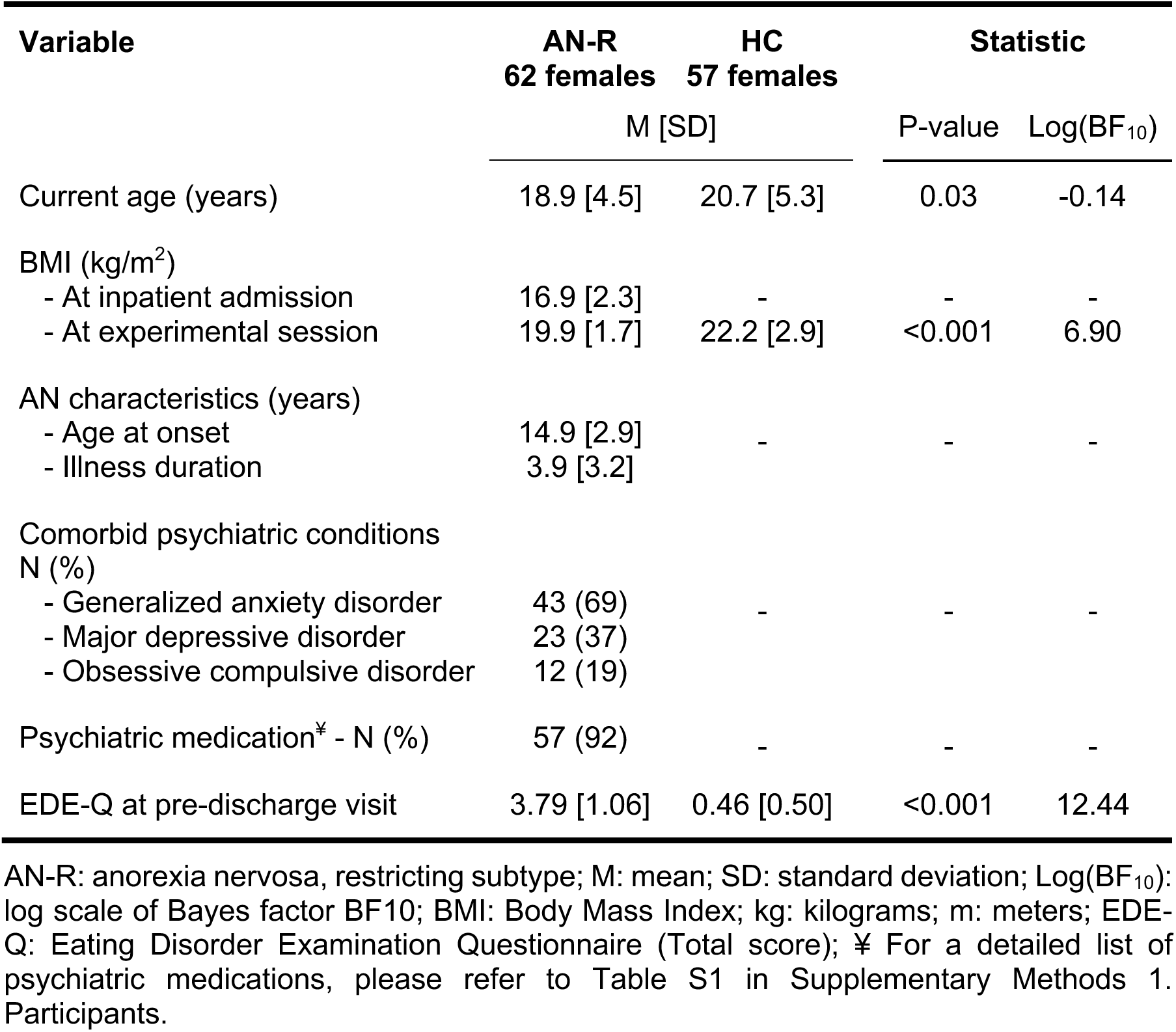
Summary of demographic and clinical information for participants with anorexia nervosa (AN) and healthy comparisons (HC) included in the cross-sectional analysis.

| Variable | AN-R<br>62 females | HC<br>57 females | Statistic |  |
| --- | --- | --- | --- | --- |
|  | M [SD] |  | P-value | Log(BF<sub>10</sub>) |
| Current age (years) | 18.9 [4.5] | 20.7 [5.3] | 0.03 | -0.14 |
| BMI (kg/m<sup>2</sup>) |  |  |  |  |
| - At inpatient admission | 16.9 [2.3] | - | - | - |
| - At experimental session | 19.9 [1.7] | 22.2 [2.9] | <0.001 | 6.90 |
| AN characteristics (years) |  |  |  |  |
| - Age at onset | 14.9 [2.9] | - | - | - |
| - Illness duration | 3.9 [3.2] |  |  |  |
| Comorbid psychiatric conditions<br>N (%) |  |  |  |  |
| - Generalized anxiety disorder | 43 (69) | - | - | - |
| - Major depressive disorder | 23 (37) |  |  |  |
| - Obsessive compulsive disorder | 12 (19) |  |  |  |
| Psychiatric medication<sup>‡</sup> - N (%) | 57 (92) | - | - | - |
| EDE-Q at pre-discharge visit | 3.79 [1.06] | 0.46 [0.50] | <0.001 | 12.44 |

AN-R: anorexia nervosa, restricting subtype; M: mean; SD: standard deviation; Log(BF<sub>10</sub>): log scale of Bayes factor BF<sub>10</sub>; BMI: Body Mass Index; kg: kilograms; m: meters; EDE-Q: Eating Disorder Examination Questionnaire (Total score); <sup>‡</sup> For a detailed list of psychiatric medications, please refer to Table S1 in Supplementary Methods 1. Participants.

### 1. Behavioral findings

Linear mixed-effects models showed significant block-by-group interactions for normalized A-prime (P=0.007, η²p=0.07; Figure 2A) and miss rate (P=0.002, η²p=0.08; Figure 2B), a main effect of group for response bias (P=0.02, η²p=0.07; Figure 2C), and no group differences in response times (RT) across blocks (P=0.13; Figure 2D). Post-hoc comparisons indicated that, during the normal stimulation condition, AN participants exhibited significantly lower interoceptive accuracy, as indicated by lower normalized A-prime values (P=0.001, Cohen’s d=-0.98; Figure 2A) and higher miss rates (P<0.001, Cohen’s d=1.02; Figure 2B); however, these differences were not observed during enhanced stimulation (normalized A-prime: P=0.87; miss rate: P=0.79). Regardless of group, normal stimulation was associated with decreased perceptual accuracy (AN: P<0.001, d=−2.22; HCs: P<0.001, d=−1.43), increased miss rates (AN: P<0.001, d=2.21; HCs: P<0.001, d=1.02), and a greater tendency not to press the button (i.e., response bias), with a significant effect in HCs (P=0.03, d=−0.61) but not in AN (P=0.10, d=−0.42).

### 2. Electroencephalogram findings

Cluster-based permutation testing did not reveal significant group differences in vibration onset-evoked event-related potential (ERP) amplitude across blocks (normal block: Monte Carlo P>0.27; enhanced block: Monte Carlo P>0.13; Figure S2 in Supplementary Results 1. Electroencephalogram findings). However, enhanced stimulations elicited significantly larger ERP amplitudes compared to normal stimulation in both groups. In the AN group, this effect occurred within a 716 millisecond (ms) window, from 244 ms to 960 ms after vibration onset (Monte Carlo P=0.005; Cohen’s d=0.54), involving the following electrodes: P3, P4, O1, O2, P8, Cz, Pz, Oz, FC1, FC2, CP1, CP2, CP6, and POz (Figure 3A). In HCs, the effect was observed within a shorter (376 ms) window, from 364 ms to 740 ms after the vibration onset (Monte Carlo P=0.03; Cohen’s d=0.53), involving a similar but not identical set of electrodes: P3, P4, O1, O2, Pz, Oz, CP1, CP2, and POz (Figure 3B).

Vibration offset-locked analyses revealed similar results. In the AN group, enhanced stimulation elicited significantly greater ERP amplitudes than normal stimulation across all electrodes, within a 1448 ms window from 128 ms to 1576 ms post-offset (Monte Carlo P=0.0009; Cohen’s d=0.63; Figure 3C). In HCs, the same effect was observed from 100 ms to 1120 ms post-offset, also across all electrodes (Monte Carlo P=0.0009; Cohen’s d=0.71; Figure 3D). We did not find significant group differences in ERP amplitude across blocks (normal block: Monte Carlo P>0.20; enhanced block: Monte Carlo P>0.34; Figure S2 in Supplementary Results 1. Electroencephalogram findings).

### 3. Computational findings

Model comparison confirmed that a model with each of the five parameters mentioned above (Model 6; see Table S4 in Supplementary Methods 4. Computational Modeling) provided the best fit to the data, with a protected exceedance probability of 1 (Table S5 in Supplementary Results 2. Model comparison and parameter recoverability). This model outperformed several alternative models that included different subsets of these and other possible parameters. Parameters in the selected model showed good recoverability, as evidenced by strong positive correlations between model parameter values used to generate simulated data and estimates for those parameter values when subsequently fitting the model to that simulated data (rs > 0.77; Ps<0.001; Table S6 in Supplementary Results 2. Model comparison and parameter recoverability). Compared to HCs, the AN group exhibited initial prior beliefs (𝑝𝑉) that more strongly favored the absence of perceiving GI signals (i.e., a stronger bias against feeling them), as indicated by 𝑝𝑉 values further below the neutral value of 0.5 (P=0.05, Cohen’s d=-0.31; Figure 4A). The AN group also exhibited a significantly greater decrease in interoceptive precision from enhanced to normal stimulation strength blocks (𝐼𝑃_*diff*_; P=0.01, Cohen’s d=0.38; Figure 4B) compared to HCs, but there was no significant group difference in overall interoceptive precision (𝐼𝑃; P=0.47). Finally, the AN group showed slower learning rates than HCs for trials with vibrations (𝜂_$_; P=0.007, Cohen’s d=-0.40) and faster learning rates than HCs for trials without vibrations (𝜂_%$_; P=0.01, Cohen’s d=0.35) (Figure 4C). The combination of group differences in initial prior beliefs (Figure 4A) and learning rate (Figure 4C) may have contributed to between-group differences in the time course of prior beliefs: based on visual inspection, the AN group showed a stronger drift in prior values toward not detecting a vibration as the task progressed (Figure 4D).

**Figure 4.**
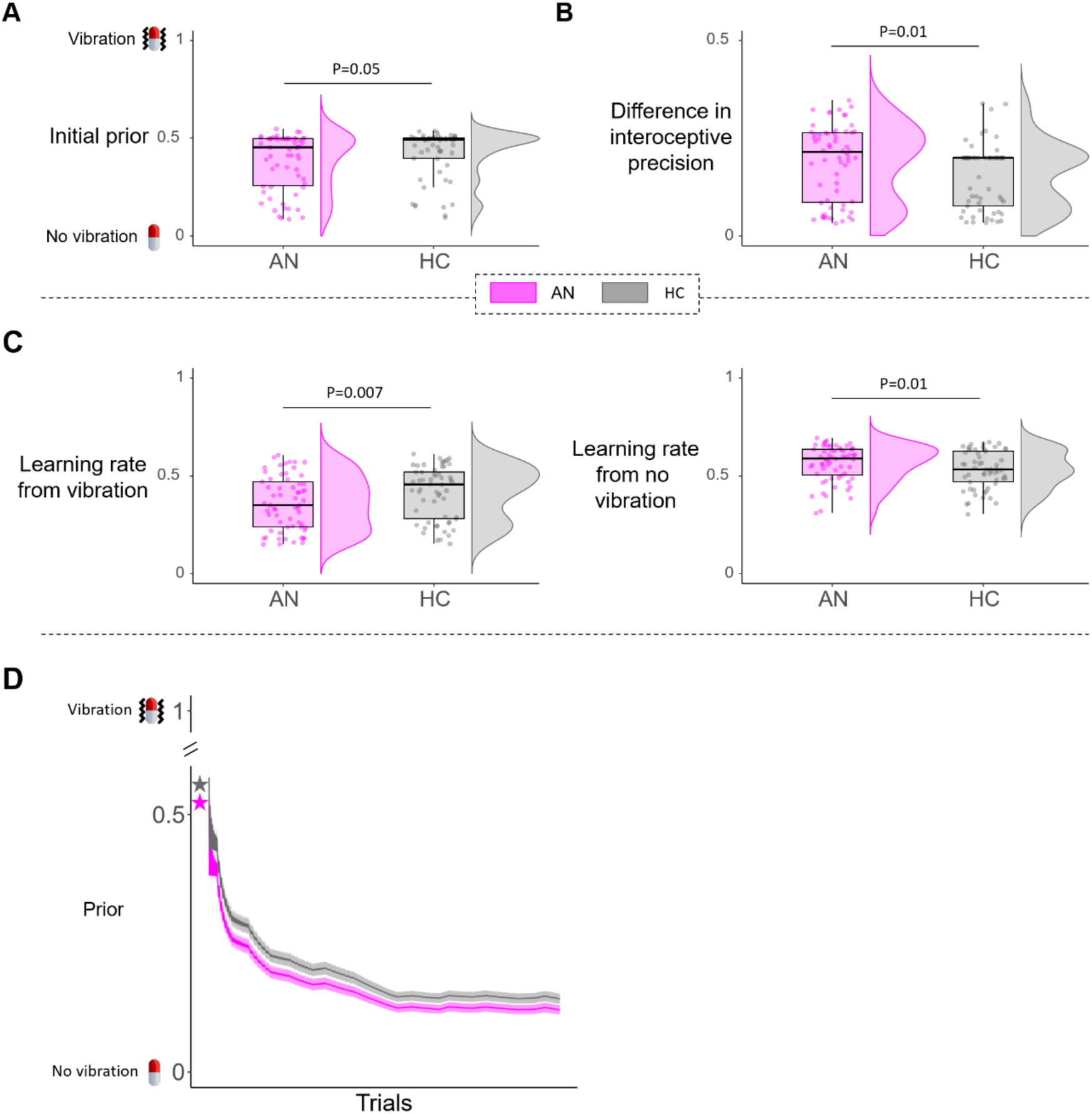
Abnormal prior beliefs, interoceptive precision differences, and learning rates in AN. Compared to HCs, the AN group exhibited stronger initial prior beliefs against perceiving vibratory gut stimulation signals **(A)**, a larger difference in interoceptive precision between normal- and enhanced-intensity stimulations **(B),** and significantly different learning rates from both vibration and no-vibration trials **(C)**. The plots display the distribution of individual data points, corresponding boxplots, and individual participant values. **(D)** Group-level time course of prior belief values across trials illustrates that the AN group showed a stronger drift below the neutral 0.5 line, indicating a stronger bias against detecting a vibration as the task progressed. Abbreviations: AN, anorexia nervosa; HC, healthy comparison.

### 4. Peripheral physiological findings

Compared to HCs, the AN group exhibited a significantly lower tonic (i.e., basal) heart rate (HR) across both the baseline period and stimulation blocks (P<0.001, η^2^p=0.37; Figure S4A). These group differences were significantly more pronounced during the stimulation blocks compared to baseline (period-by-group interaction effect: P=0.05, η^2^p=0.03). However, no significant group differences were observed for phasic HR activity (i.e., rapid responses specific to the 3-second vibration period; P=0.72) or heart rate variability (P=0.17), as measured using the standard deviation of R–R intervals (SDNN) (Figure S4B-C). Further, no significant group differences were observed for electrogastrogram (EGG) responses (Figures 4D-E-F-G), including the absolute power across four gastric frequency ranges: total power (0.5–11 cpm; P=0.85), bradygastria (0.5–2.25 cpm; P=0.88), normogastria (2.5–3.5 cpm;

P=0.86), and tachygastria (3.75–9.75 cpm; P=0.47). Finally, no group differences were observed in skin conductance response (SCR), specifically in the maximum value of phasic activity (P=0.97; Figure S4H).

### 5. Longitudinal findings

At the 6-month follow-up, 16 AN individuals were classified as fully relapsed, while 38 were classified under another AN-related condition including partial recovery, full remission, partial remission, or partial relapse (see Supplementary Methods 6 and Figure S5). 6-month data were missing for 8 AN individuals due to dropout.

Relapse outcome at 6 months was successfully predicted by experimental session measures. Specifically, initial prior beliefs (𝑝𝑉) (Odds Ratio (OR)=3.82; P=0.05) and response bias in the normal block (OR=5.37; P=0.04) predicted relapse, such that AN individuals with stronger initial prior beliefs against perceiving GI signals, as reflected by lower 𝑝𝑉 values, were more likely to be fully relapsed at six months. Similarly, AN individuals with more positive response bias values, meaning they were less likely to press the button during normal stimulations, were also more likely to be in full relapse at six months. Finally, eating disorder symptom severity at 6 months, indexed by EDE-Q scores, were also significantly predicted by several experimental session measures, including miss rate (P=0.05; adjusted R squared (R^2^)=0.08), difference in 𝐼𝑃 between normal and enhanced stimulations (𝐼𝑃_*diff*_; P=0.004; R^2^=0.16), and initial prior beliefs (𝑝𝑉; P=0.05; R^2^=0.09) (Supplementary Results 4. Longitudinal findings).

### 6. Multilevel correlation findings

The average ERP amplitude (avERP), derived from the spatiotemporal window identified as the EEG marker of gut mechanosensation (see Figures 3A-B), was significantly correlated with perceptual accuracy measures during the normal stimulation block across diagnostic groups (AN and HCs). Specifically, avERP amplitude was positively correlated with normalized A-prime (AN: *r*=0.70, p<0.001; HC: *r*=0.49, p<0.05) and negatively correlated with miss rate (AN: *r*=-0.66, p<0.001; HC: *r*=-0.57, p<0.001).

Additionally, avERP amplitude showed significant associations with several computational parameters: 𝐼𝑃_*diff*_ in the AN group only (AN: *r*=-0.62, p<0.001; HC: *r*=-0.37, p=0.69)), the learning rate for vibration trials in both groups (𝜂_$_; AN: *r*=0.58, p<0.001; HC: *r*=0.49, p<0.01), and the learning rate for no-vibration trials in both groups (𝜂_%$_; AN: *r*=-0.60, p<0.001; HC: *r*=-0.46, p<0.05).

Across all of these variables, correlations were consistently larger in the AN group, with higher absolute correlation coefficients and greater statistical significance compared to HCs (Supplementary Results 5. Multilevel correlation findings). Furthermore, avERP amplitude was significantly and inversely associated with response times during the normal stimulation block in both groups (AN: *r*=-0.60, p<0.01; HC: *r*=-0.68, p<0.001).

No significant correlations were observed between avERP amplitude and response bias (AN: *r*=-0.01, p=1; HC: *r*=-0.16, p=1), prior beliefs (𝑝𝑉; AN: *r*=0.26, p=1; HC: *r*=0.27, p=1), or interoceptive precision (𝐼𝑃; AN: *r*=0.27, p=1; HC: *r*=0.11, p=1). Likewise, avERP was not significantly associated with any behavioral or computational measures during the enhanced stimulation block (Supplementary Results 5. Multilevel correlation findings).

### 7. Self-report findings

#### 7.1.1. Interoceptive intensity

For stomach/digestive sensations, intensity increased during the task (P<0.001; η²p=0.49), but surprisingly, it did not differ by group (P=0.73) (Figure 5A). For breathing sensations, there was a main effect of group (P<0.001, η²p=0.14; Figure 5B), with AN individuals reporting greater breathing sensation intensity than HCs both before (P=0.007, Cohen’s d=0.65) and during the task (P<0.001, Cohen’s d=1.03). For heartbeat sensations, the group-by-time interaction was significant (P=0.004, η²p=0.07; Figure 5C). Post-hoc comparisons indicated that AN participants reported a significant increase from before to during the task (P<0.001, Cohen’s d=0.86), whereas HCs did not (P=0.61). For muscle tension, there was a main effect of group (P<0.001, η²p=0.13; Figure 5D), with AN individuals reporting greater muscle tension than HCs both before (P<0.001, Cohen’s d=1.03) and during the task (P=0.01, Cohen’s d=0.70). Pre-task stomach/digestive intensity ratings predicted EDE-Q global symptom scores at 6 months (P = 0.01; R^2^=0.13); no other intensity measure predicted relapse (Supplementary Results 6).

**Figure 5.**
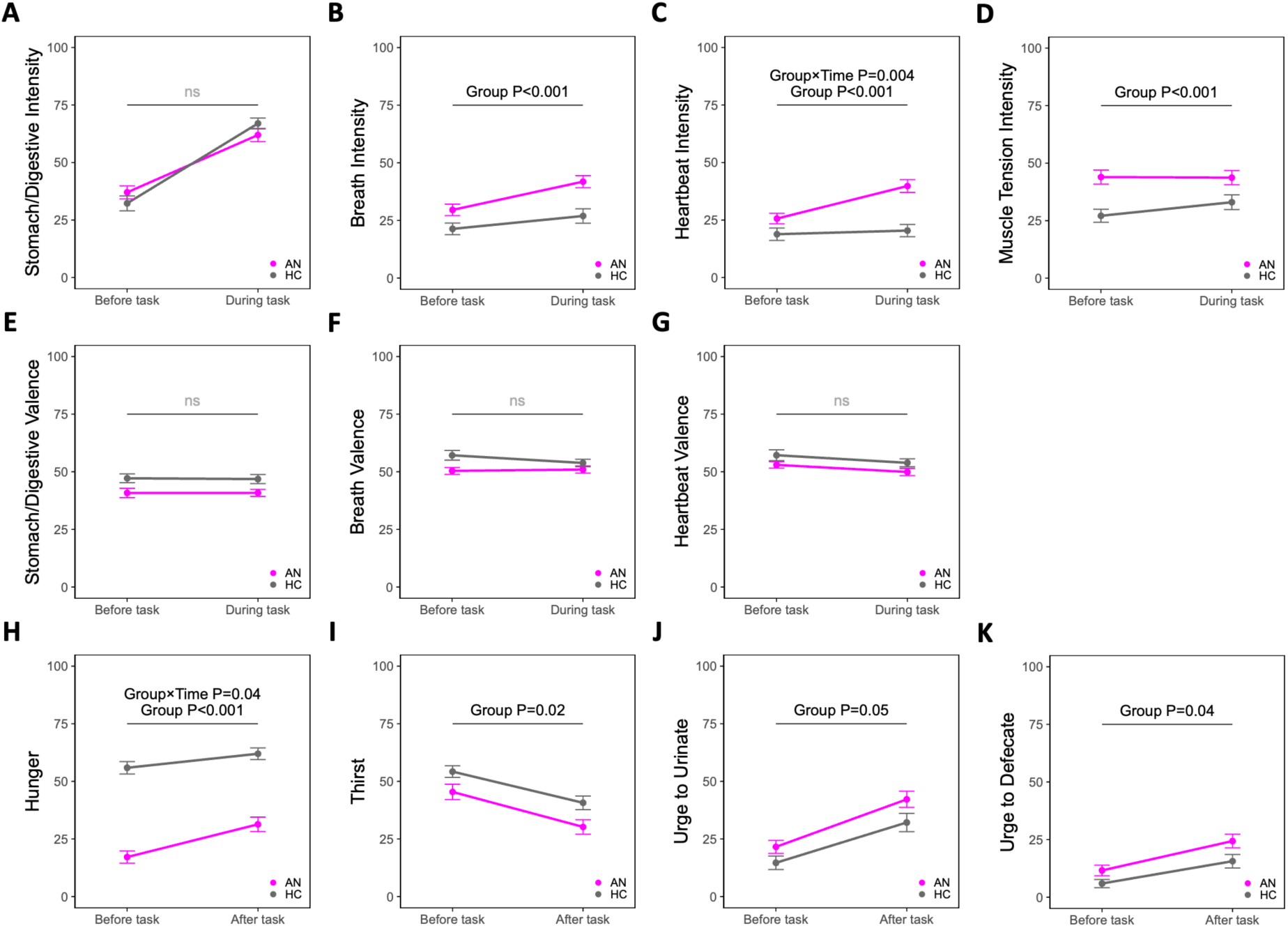
Retrospective ratings of interoceptive sensations and homeostatic urges before, during, and after vibratory gut stimulation. **(A)** Stomach/digestive intensity: increased during the task in both groups, with no group differences. **(B)** Breathing intensity: higher in individuals with AN than HCs both before and during the task. **(C)** Heartbeat intensity: group by time interaction, with an increase during the task in AN but no change in HCs. **(D)** Muscle tension: higher in AN than HC both before and during the task. **(E–G)** Valence: marginal group differences for stomach/digestive and breathing; none for heartbeat. **(H)** Hunger: group by time interaction, with a larger task-associated increases in the AN group. **(I)** Thirst: decreased during the task in both groups and was consistently lower in the AN group than HC. **(J–K)** Urges to urinate/defecate: increased from pre to post in both groups and were higher in AN than in HC. **Note**: Main effects of time were observed for Stomach, Breath, Heartbeat intensity (A–C), Heartbeat valence (G), and Hunger, Thirst, Urination and Defecation urge (H–K) but are not displayed (see results for details). Muscle tension valence was not assessed. Abbreviations: AN, anorexia nervosa; HC, healthy comparison.

#### 7.1.2. Interoceptive valence

Group differences in valence were marginal for stomach/digestive (P=0.06; Figure 5E) and breathing sensations (P=0.07; Figure 5F), and non-significant for heartbeat sensations (P=0.24; Figure 5G). No significant effects of time or group-by-time interactions were observed for any of the valence measures. However, in the AN group, relapse status at 6 months was predicted by the valence of stomach sensations: higher stomach unpleasantness during the vibrating capsule task was associated with greater odds of full relapse (OR=5.73, P=0.03).

Eating disorder symptom severity at 6 months was also predicted by stomach valence ratings before (P=0.02, R²=0.11) and during the task (P=0.02, R²=0.11; Supplementary Results 6).

#### 7.1.3. Homeostatic interoceptive urges

For hunger, linear mixed-effects models showed a significant group-by-time interaction (P=0.04, η²p=0.04; Figure 5H). Post-hoc comparisons indicated that task-induced increases in hunger were larger in AN individuals (P<0.001, Cohen’s d=0.94) than in HCs (P=0.03, Cohen’s d=0.40). Thirst decreased after the task in both groups (P<0.001; η²p=0.28; Figure 5I) and was consistently lower in the AN group than in HCs (P<0.001; η²p=0.05). The urge to urinate increased after the task in both groups (P<0.001; η²p=0.36; Figure 5J) and was higher in AN individuals than in HCs (P=0.05; η²p=0.03). Urge to defecate likewise increased in both groups after the task (P<0.001; η²p=0.17; Figure 5K) and was higher in the AN group than in HCs (P=0.04; η²p=0.04). However, none of these items were predictive of relapse outcome or EDE-Q score at 6 months (Supplementary Results 6).

## DISCUSSION

In this study, we applied a clinically scalable mechanosensory stimulation paradigm to probe GI interoception in weight-restored females with restrictive AN, revealing pervasive disruptions across behavioral and computational levels. Compared to matched healthy comparisons, individuals with AN showed reduced accuracy in detecting normal-intensity gut sensations, accompanied by higher miss rates and lower normalized A-prime scores. Computational modeling revealed that AN participants held stronger prior beliefs against feeling gut sensations and demonstrated asymmetric learning—specifically, diminished updating from vibration presence and exaggerated updating its absence—resulting in a progressive amplification of biased priors over time. While overall interoceptive precision did not differ between groups, individuals with AN displayed greater sensitivity to changes in stimulation magnitude reflected in greater reductions in precision between enhanced and normal stimulation conditions. Although capsule-evoked ERP amplitudes did not differ overall between groups, individuals with AN showed earlier and more prolonged ERP differences between conditions, along with stronger correlations between neural signals and behavioral/computational measures during normal-intensity stimulation. Notably, several task-derived markers, including initial prior beliefs and response bias, predicted clinical relapse at six-months, identifying these as candidate prognostic biomarkers. Self-report ratings also held prognostic value: greater stomach unpleasantness during stimulation predicted full relapse, while perceived stomach intensity and valence correlated with future symptom severity. Lastly, the AN group exhibited a larger increase in hunger during stimulation, suggesting altered homeostatic interoceptive responses.

These findings offer direct empirical support for disrupted GI interoception in weight-restored individuals with AN, evidenced by reduced accuracy in detecting mechanosensory gut signals. They extend theoretical models positing interoceptive dysfunction as a core mechanism in AN pathophysiology^14^, while addressing an important gap in the literature predominantly focused on cardiac interoception^13^. Despite the GI tract’s essential role in regulating satiety and visceral discomfort, features central to disordered eating, studies of GI interoception in AN have been limited by methodological barriers. Invasive approaches, such as esophageal balloon inflation^43,44^, are ill-suited for psychiatric settings, while tools such as water load testing^45,46^ are limited to single-trial assessments that preclude assessment of dynamic learning mechanisms. The vibrating capsule approach overcomes these limitations, offering a non-invasive, repeatable, and developmentally appropriate method for probing gut-brain signaling across adolescence and young adulthood, when AN often emerges. Although our self-report findings on cardiac sensations replicate prior evidence of altered cardiac interoception in AN^47–51^, the current results underscore the need to broaden mechanistic investigation beyond the cardiac axis to more symptom-relevant domains of the viscera.

Computational modeling revealed altered interoceptive inference in AN, characterized by stronger prior beliefs against detecting gut sensations and maladaptive learning dynamics. These findings align with Bayesian computational frameworks of interoceptive psychopathology suggesting that excessive reliance on prior expectations, or hyperprecise priors, can distort bodily perception across psychiatric disorders^17,20^. In AN, such priors may suppress ascending visceral signals, contributing to misinterpreted or blunted satiety cues. This mechanism may also apply to hunger and thirst cues, supporting the ability of individuals with AN to ignore hunger/thirst signals. Supporting this possibility, individuals with AN updated less from true vibrations and more from their absence, a learning asymmetry that may promote escalating rigidity in belief systems. This dynamic parallels known cognitive inflexibility in AN^52^ and suggests a mechanistic link between perceptual inference and executive dysfunction. Future studies should investigate whether this rigidity reflects interoceptive-specific learning or broader impairments in inhibitory cognitive control^53^. From a normative perspective, such asymmetry in learning may arise from metacognitive beliefs, learned over longer timescales, that internal states are unreliable or unchangeable^54^. More broadly, our findings extend prior computational work in visual and cardiac domains^19,55^ highlighting that disruptions in interoceptive inference persist after weight restoration. These results underscore recent clarion calls to move beyond nutritional rehabilitation alone^56^, and suggest that computational interoceptive markers could inform brain-gut psychotherapies, such as GI-focused cognitive-behavioral therapy, aimed at recalibrating maladaptive priors and promoting flexible learning from visceral input^57^.

From a neurophysiological perspective, this study provides the first evidence linking capsule-evoked ERPs to behavioral and computational markers of GI interoception in AN. Although overall ERP amplitudes did not differ by group, clear onset- and offset-locked responses emerged across participants, with amplitudes strongly modulated by stimulation intensity. These results confirm that the capsule reliably engages cortical activity involved in visceral signal processing, underscoring its value as a sensitive tool for probing gut-brain communication. Importantly, ERP amplitudes during normal-intensity stimulation were closely associated with perceptual accuracy and learning rates, particularly in the AN group. This suggests that gastric ERPs may reflect both bottom-up sensitivity to mechanosensory input, and top-down flexibility in belief updating, echoing the functional interpretations of heartbeat-evoked potentials^58,59^ and P300 components^60,61^. Interestingly, ERP amplitudes correlated positively with perceptual accuracy, in contrast to cardiac interoceptive prediction error signals, which are often negatively correlated with dysfunction^17,62–64^. This raises the possibility that GI interoceptive deficits in AN may involve compensatory processes or active suppression of sensory input, rather than overt prediction-error amplification alone. Beyond amplitude, AN individuals also showed earlier onset and prolonged duration of ERP responses. Although the functional significance of this expanded temporal envelope remains unclear, it may reflect heightened salience attribution or sustained attentional engagement with visceral stimuli, as suggested by prior ERP duration studies^65,66^. Such prolonged engagement could contribute to persistent monitoring of bodily signals in AN, reinforcing symptom expression and potentially increasing relapse risk.

Multimodal physiological assessment revealed selectively altered cardiac function in AN, specifically lower tonic heart rate during rest and stimulation, consistent with parasympathetic dominance following weight restoration^67,68^. In contrast to prior reports, we observed no significant group differences in heart rate variability, electrogastrogram, or skin conductance responses. These discrepancies may reflect methodological differences, including the short-term, task-based nature of our recordings versus the extended monitoring protocols (e.g., 24-hour) in prior studies^67,68^, which may capture different aspects of autonomic regulation over time^33^. Alternatively, normalization of autonomic indices during inpatient recovery may have attenuated group differences^69^. The absence of EGG or SCR abnormalities, despite robust behavioral and computational group differences, contrasts with heterogeneous samples in prior literature^46,67,70,71^, highlighting the specificity of interoceptive disruption in perceptual and inferential domains rather than global autonomic dysfunction. These results suggest that clinically relevant interoceptive alterations in AN may not be fully captured by conventional physiological metrics alone, underscoring the added value of integrated perceptual and modeling-based approaches.

Longitudinal analyses revealed that both behavioral and computational markers of interoceptive processing prospectively predicted clinical relapse in AN. Specifically, greater response bias and stronger prior beliefs against perceiving gut signals were associated with full relapse at six months, while miss rate and interoceptive precision shifts predicted future eating disorder symptom severity. These objective, task-derived markers offer a promising complement to self-report-based assessments^72^, which may be compromised in AN by limited introspective ability or alexithymia^73,74^. Intriguingly, task-evoked changes in subjective experience, particularly intensity and valence of stomach sensations, also predicted relapse and symptom outcomes. While stomach unpleasantness may signal heightened visceral threat sensitivity, the larger increase in hunger following mechanosensory stimulation in AN raises the possibility of preserved but dormant homeostatic signaling. This unforeseen observation suggests that controlled gastric mechanosensory stimulation may have potential as a therapeutic tool to reconnect patients with internal hunger cues, especially in those who experience profound disconnection from appetite. If replicated, such bottom-up stimulation paradigms could support meal initiation and augment refeeding efforts during early recovery. Together, these findings support the use of interoceptive challenge tasks not only for stratifying relapse risk, but also for identifying novel targets to enhance treatment engagement in AN.

Self-report ratings complemented task-based interoceptive measures, revealing modality-specific alterations in AN. Individuals with AN reported greater breathing, heartbeat, and muscle tension intensity across timepoints, consistent with prior evidence of heightened interoceptive attention to cardiopulmonary signals^12,75^. In contrast, perceived stomach sensation intensity increased with stimulation but did not differ by group, despite clear behavioral impairments in AN. This dissociation highlights the limited utility of subjective ratings alone in capturing visceral processing abnormalities and underscores the need for concurrent behavioral and computational assessment. However, stomach unpleasantness predicted relapse, and pre-task stomach intensity ratings were associated with future symptom severity, suggesting that task-evoked self-report may still hold prognostic value when embedded within interoceptive perturbation paradigms.

This study has several limitations. First, the sample included only weight-restored females with restrictive AN, limiting generalizability to males, individuals with binge-purge presentations, and those who are acutely underweight. Second, all participants were recruited from a single inpatient program, which may reduce applicability to more heterogeneous outpatient or community samples. Third, while the vibrating capsule offers advantages in scalability and non-invasiveness, it targets primarily gastric and proximal small intestinal mechanoreceptors, leaving other visceral modalities (e.g., chemosensory, hormonal) unaddressed. Additionally, physiological recordings were brief and task-bound, potentially missing subtler or slower-evolving autonomic patterns detectable via extended monitoring. Finally, although the six-month follow-up allowed for relapse prediction, longer-term trajectories of remission and recurrence (e.g., 1-year and beyond^4^) remain unexplored. Despite these limitations, the integration of behavioral, neural, and computational approaches in this study provides a novel and mechanistically rich framework for understanding and potentially intervening upon disordered interoception in AN.

## CONCLUSION

This study demonstrates that weight-restored individuals with AN exhibit pronounced disruptions in gastrointestinal interoception, including reduced interoceptive accuracy, maladaptive prior beliefs, and inflexible interoceptive learning. These abnormalities were identified across behavioral, neural, and computational domains, and several markers prospectively predicted relapse, underscoring their potential as clinically actionable biomarkers. By combining a non-invasive, scalable mechanosensory probe with computational modeling and EEG, this work offers a mechanistic framework for stratifying risk and tracking interoceptive function in treatment. More broadly, the findings support the development of brain–gut interventions aimed at recalibrating bodily beliefs, restoring appetite awareness, and improving long-term outcomes in eating disorders.

## Data Availability

All data produced in the present study are available upon reasonable request to the authors.

## Acknowledgments.

Alexa Morton and Rayus Kuplicki provided assistance with data collection and physiological data processing. Preliminary results from this study were presented at the 2023 and 2024 annual meetings of the American College of Neuropsychopharmacology and at the 2025 ECNP Workshop on Applied Neuroscience.

## Author Contributions

Concept and design: Khalsa. Acquisition of data: Mink, Verdonk, Khalsa. Processing of data: Verdonk, Mink, Mayeli. Analysis or interpretation of data: Verdonk, Keller, Choquette, Smith, Khalsa. Drafting of the manuscript: Verdonk, Khalsa. Critical revision of the manuscript for important intellectual content: all authors. Statistical analysis: Verdonk, Khalsa. Obtained funding: Khalsa. Administrative, technical, or material support: Khalsa, Paulus. Supervision: Khalsa, Smith.

