## Supplement for "Altered Gastrointestinal Interoception in Anorexia Nervosa Predicts Relapse"

**Table of contents**

**Supplementary Methods 1.** Participants..…………………………………….…………….…..……..Page 3
**Supplementary Methods 2.** Experimental protocol..……………………………………...……..Page 7
**Supplementary Methods 3.** Behavioral measures and self-report measures……..……...Page 9
**Supplementary Methods 4.** Computational modeling..…....…………………………..….…..…..Page 10
**Supplementary Methods 5.** Processing of peripheral physiological data………………....Page 14
**Supplementary Methods 6.** Illness status definitions…………………………….….………..Page 15
**Supplementary Methods 7.** Statistical analysis…......…………………………….…..………Page 17
**Supplementary Results 1.** Electroencephalogram findings……......……………….…………Page 19
**Supplementary Results 2.** Model comparison and parameter recoverability……………..Page 21
**Supplementary Results 3.** Peripheral physiological findings.....................…….…..………..Page 23
**Supplementary Results 4.** Longitudinal findings..........…...…………………………..………..Page 24
**Supplementary Results 5.** Multilevel correlation findings.....……………………….…..……..Page 27
**Supplementary Results 6. Self-report findings**.....……………………..…………….…..Page 29

**Supplementary Methods 1.** Participants

1. **Participant flow**

One hundred eighty-seven participants provided informed written consent and were assessed for eligibility. Thirty-two participants were excluded due to exclusionary medical conditions (see Section 1: Exclusion Criteria below), and an additional nine participants withdrew prior to the intervention for personal reasons. Of the 146 participants allocated to the intervention, 78 individuals were diagnosed with anorexia nervosa (AN) restrictive subtype and 68 individuals were defined as healthy comparisons (HCs). One AN participant and one HC participant withdrew before any data collection due an inability to swallow the capsule. After completion of the intervention, data related to 25 participants were discarded for the following reasons: inadequate behavioral response registration (n=4), corrupted behavioral data file (n=1), corrupted electroencephalography (EEG) data files (n=5), low quality Event Related Potential (ERP) signal (n=9), and low-quality capsule signal (n=6). For follow-up data, 6-month results from 8 individuals with AN were missing due to passive refusals (not responding to remote follow-up requests) (Figure S1). The data were collected from August 15, 2020 to December 05, 2024 at the Laureate Institute for Brain research (Tulsa, Oklahoma).

2. **Exclusion criteria**

Participants were excluded from the study if they had a diagnosis of a psychosis-spectrum disorder or bipolar disorder, or endorsed severe behavioral disturbances such as active suicidal ideation, recent self-harm behavior, active purging, or a severe history of purging. Pregnancy or lactation was also exclusionary. Gastrointestinal-related exclusions included diagnoses such as inflammatory bowel disease, gastroparesis, complicated diverticular disease, intestinal obstruction, mega-rectum or colon, congenital anorectal malformation, and rectal prolapse (Table S2). Participants with a history of intestinal resection (excluding appendectomy, cholecystectomy, or inguinal hernia repair), bariatric surgery, or other structural abnormalities that could impact gastrointestinal transit were excluded. Additionally, individuals with esophageal disorders (such as Zenker's diverticulum, dysphagia, Barrett's esophagus, achalasia, or eosinophilic esophagitis) were not eligible. Cardiovascular exclusion criteria included orthostatic hypotension and bradycardia defined as a heart rate below 40 beats per minute. Pain disorders were also exclusionary. Finally, chronic use of non-steroidal anti-inflammatory drugs, defined as taking full dose NSAIDs more than three times a week for at least six months, and regular use of medications known to significantly alter gastrointestinal motility—such as high-dose prokinetics (e.g., metoclopramide, erythromycin, senna, prucalopride), anti-Parkinsonian agents, opioids, calcium-channel blockers, or frequent use of enemas—were grounds for exclusion.

**Table S1.** Diagnostic comorbidities, psychotropic medication status, and race of anorexia nervosa (AN) and healthy comparison (HC) participants.

| | AN (n=62 females) | HC (n=57 females) |
|---|---|---|
| **Comorbid Diagnoses** - N (%) | | |
| Generalized anxiety disorder | 43 (69) | 0 |
| Major depressive disorder | 23 (37) | 0 |
| Obsessive–compulsive disorder | 12 (19) | 0 |
| **Psychotropic medication** - N (%) | 57 (92) | |
| Adrenergic antagonist[1] | 2 (3) | |
| Dopamine modulator[2] | 19 (31) | |
| GABA modulator[3] | 3 (5) | |
| Glutamate modulator[4] | 5 (8) | |
| Histamine antagonist[5] | 23 (37) | 0 |
| Lithium enzyme modulator[6] | 1 (2) | |
| Melatonin agonist[7] | 21 (34) | |
| Norepinephrine modulator[8] | 19 (31) | |
| Opioid antagonist[9] | 1 (2) | |
| Serotonin modulator[10] | 57 (92) | |
| **Race** | | |
| White | 56 (90) | 42 (74) |
| Asian | 2 (3) | 9 (16) |
| Native American | 3 (5) | 1 (2) |
| Middle Eastern | 1 (2) | 0 |
| Black | 0 | 3 (5) |
| Other | 0 | 2 (3) |

M: mean; SD: standard deviation; N: number of participants; %: proportion of participants
[1]propanolol; [2]amantadine, aripiprazole, brexpiprazole, bupropion, cariprazine, lurasidone, olanzapine, risperidone; [3]lorazepam, melatonin-GABA-valerian supplement; [4]gabapentin, lamotrigine; [5]cetirizine, diphenhydramine, doxepin, fexofenadine, hydroxyzine, levocetirizine, loratadine, quetiapine ; [6]lithium carbonate; [7]melatonin supplement, melatonin-GABA-valerian supplement; [8]atomoxetine, amitriptyline, bupropion, desvenlafaxine, duloxetine, mirtazapine, risperidone, venlafaxine; [9]naltrexone; [10]amitriptyline, aripiprazole, brexpiprazole, buspirone, cariprazine, desvenlafaxine, duloxetine, escitalopram, fluoxetine, fluvoxamine, lurasidone, mirtazapine, olanzapine, risperidone, sertraline, trazodone, venlafaxine, vilazodone.

**Table S2.** Exclusion criteria.

**Exclusionary condition**

- Psychosis spectrum disorder or bipolar disorder
- Active suicidal ideation, engagement in self-harming behaviors, ongoing purging behaviors, or a severe history of purging
- Pregnancy and lactation
- Significant gastrointestinal disorder, including any form of inflammatory bowel disease or gastrointestinal malignancy (celiac disease is accepted if the subject has been treated and is in remission)
- Complicated/obstructive diverticular disease
- Gastroparesis
- Mega-rectum or colon, congenital anorectal malformation, or clinically significant rectocele or rectal prolapse
- Intestinal or colonic obstruction, or suspected intestinal obstruction
- Intestinal resection (with an exception for appendectomy, cholecystectomy and inguinal hernia repair), bariatric surgery or evidence of any structural abnormality of the gastrointestinal tract that might affect transit
- Zenker’s diverticulum, dysphagia, Barrett's esophagus, esophageal stricture or achalasia, transesophageal fistula, or eosinophilic esophagitis.
- Clinical evidence (as judged by the investigator) of respiratory, cardiovascular, renal, hepatic, biliary, endocrine, or neurologic disease
- Orthostatic hypotension
- Gastrointestinal bleed within the last 3 months
- Pelvic floor dysfunction/defecatory disorder
- Bradycardia with resting heart rate less than 40 bpm
- Pain disorder
- Known allergy to soybeans, beeswax, or calcium carbonate

**Exclusionary medication**

- Chronic use of non-steroidal anti-inflammatory drugs (NSAIDs), defined as taking full dose NSAIDs more than three times a week for at least six months. Those taking cardiac (i.e., low) doses of aspirin may be enrolled
- Regular use of any of the following medications or procedures: Medications that may substantially affect intestinal motility, prokinetics at high doses (metoclopramide, erythromycin, senna, prucalopride), anti-Parkinsonian medications, opiates, opioids, calcium-channel blockers, enemas

**Figure S1 – CONSORT flow diagram for the study**. AN: anorexia nervosa; EEG: electroencephalography; ERP: Event Related Potential.

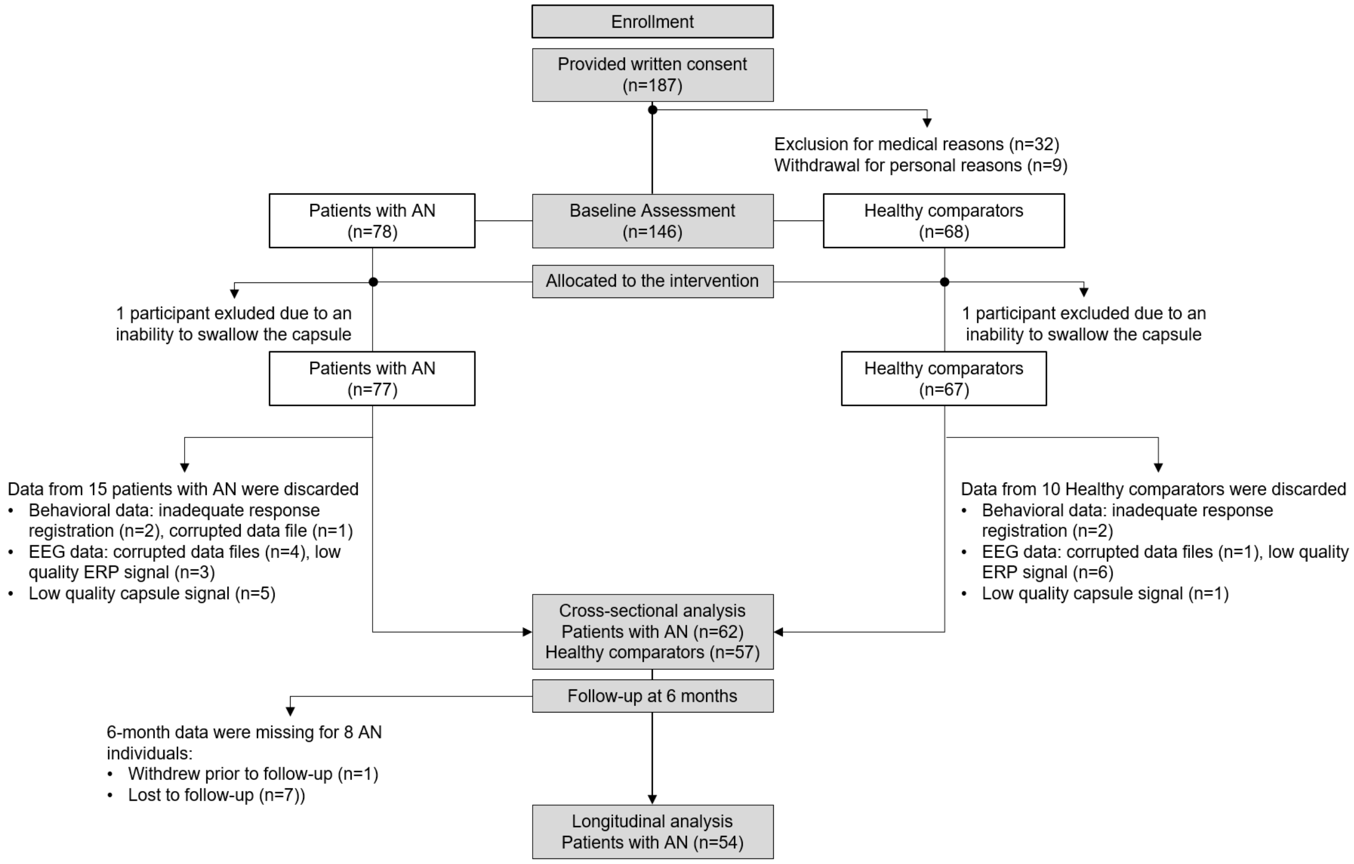

**Supplementary Methods 2.** Experimental protocol

### 1. Meal protocol

Participants were instructed to fast from both food and liquids (excluding water) for at least three hours prior to the capsule session and were informed they would not be permitted to eat or drink during the visit. This fasting protocol was implemented to allow gastric emptying to occur and to reduce baseline gastrointestinal activity prior to capsule-related measurements. For participants with AN, fasting and scheduling were coordinated with their inpatient treatment program. AN participants ate lunch at 12:00 PM on the unit, attended the study session from 2:00 to 5:00 PM (with the capsule swallow task occurring at approximately 4:00 PM), and returned to the inpatient unit following the appointment to eat dinner at 5:00 PM in accordance with their meal plan. Thus, AN participants were in a pre-meal phase during capsule stimulations, with the expectation that they would eat a meal upon completion of the experiment. HC participants followed the same fasting protocol but were scheduled independently.

### 2. Vibrating capsule

The Vibrant capsule, developed by Vibrant Ltd, has received marketing authorization by the Food and Drug Administration for the treatment of adults with chronic idiopathic constipation by delivering mechanical stimulation directly to the colon. It is an orally administered, non-biodegradable device that can be wirelessly activated using a specialized activation unit (see Figure 1 in the main text). The Vibrant capsule is classified as a non-significant risk (NSR) device, and its safety has been validated in studies involving both healthy volunteers (Ron, Halpern et al. 2015, Mayeli, Al Zoubi et al. 2023) and patients with chronic constipation (Nelson, Camilleri et al. 2017, Rao, Lembo et al. 2020).

### 3. Masking procedure

To manage participants' expectations, they were informed that two different versions of the Vibrant capsule were being tested and that they would be randomly assigned to one of three study groups: two involving vibrating capsules (modes A or B) or a non-vibrating placebo capsule. Participants were also told that neither they nor the experimenter would know whether any stimulation would occur. In reality, all participants received a vibrating capsule, making this a single-blinded study. All participants were instructed to fast for three hours before the study to ensure an empty stomach, based on the rationale that a healthy person without GI disorders would likely have an empty stomach three hours after eating.

### 4. Mechanosensory stimulation

Mechanosensory stimulations began shortly after participants ingested the activated Vibrant capsule, which was swallowed with approximately 240 ml of water while they sat comfortably in a chair. During the session, they were instructed to focus their attention on abdomen sensations while fixating on a cross on a monitor positioned about 60 cm away. They used their dominant hand to press a button when they detected the capsule’s vibrations and released it once the sensation ended. Stimulations commenced approximately three minutes after the capsule’s activation. Participants remained seated throughout the experiment to reduce motion artifacts in the EEG and EGG recordings. They were advised to keep their non-dominant hand on their lap and avoid touching their abdomen, while a research assistant seated behind them in the room monitored their alertness and adherence to instructions. Each participant underwent two blocks of stimulation (normal and enhanced) in a counterbalanced order, with each block delivering 60 stimulations over a 13-minute period. A 4-minute break separated the blocks, resulting in a total stimulation period of approximately 33 minutes.

#### 5. Vibration detection

To accurately record the timing of each vibration, a digital stethoscope (Thinklabs Inc.) was affixed to the lower right quadrant of the abdomen using a Tegaderm patch (15 x 20 cm). The stethoscope signal was continuously recorded throughout the experiment at a sampling rate of 1000 Hz. Custom scripts in Matlab 2021a (Mathworks®) were developed to identify the onset and offset of each vibration. The timing of each vibration was then manually verified and adjusted if necessary. Participants whose vibration amplitudes could not be reliably detected due to technical issues were excluded from the analysis (Figure S1).

### Supplementary Methods 3. Behavioral measures and self-report measures

1. Behavioral measures

We calculated perceptual accuracy for each participant and experimental block using a non-parametric signal detection measure analogous to $d'$, which is suitable for conditions with a lower number of trials (Grier 1971, Stanislaw and Todorov 1999). Specifically, the perceptual accuracy measure called $A'$ (A prime) was computed as (Equation 1):

$$\mathrm{A}' = \begin{cases} 0.5 + \dfrac{(H-F)(1+H-F)}{4H(1-F)} & when\ H \geq F \\ 0.5 - \dfrac{(F-H)(1+F-H)}{4F(1-H)} & when\ H < F \end{cases} \tag{1}$$

where $H$ represents the hit rate and $F$ denotes the false-alarm rate.

The $A'$ scores were then normalized using Equation 2, yielding values between 0 and $\pi$.

$$\text{Normalized A Prime} = 2\sin^{-1}\left(\sqrt{A'}\right) \tag{2}$$

We also calculated response bias, which captures the overall tendency to press the button. Positive values (ranging from 0 to 1) indicate a tendency not to press the button, whereas negative values (ranging from -1 to 0) indicate a tendency to press it (Grier 1971, Stanislaw and Todorov 1999). Response bias was computed as follows (Equation 3):

$$\mathrm{B}'' = \begin{cases} \dfrac{H(1-H)-F(1-F)}{H(1-H)+F(1-F)} & when\ H \geq F \\ \dfrac{F(1-F)-H(1-H)}{F(1-F)+H(1-H)} & when\ H < F \end{cases} \tag{3}$$

where $H$ represents the hit rate and $F$ denotes the false-alarm rate.

Additional behavioral measures included the miss rate and response time, the latter defined as the difference between the onset of the vibration and the participant's button press, indicating their perception of the sensation.

~~1.~~2. Self-report measures

Before and after the task, participants completed visual analog scales (0–100). Retrospective items assessed intensity and valence for the stomach/digestive system, breath, and heartbeat; muscle tension was assessed for intensity only. State items assessed current hunger, thirst, and urges to urinate/defecate.

Pre-task stems referenced the past hour (e.g., "Over the past hour, how intensely did you feel your [stomach/digestive system/breath/heartbeat]?"; "…how pleasant or unpleasant were your [same]?"), and post-task stems referenced the stimulation period (e.g., "During the capsule stimulation, how intensely…?"; "…how pleasant or unpleasant…?"). For muscle tension, the pre-task item used a current ("right now") stem and the post-task item used a retrospective ("during capsule stimulation") stem. State items used identical wording at both time points (e.g., "How [hungry/thirsty/much of an urge to urinate/defecate] do you currently feel?").

Scale anchors were: intensity/state 0 = "Not at all/None," 100 = "Extremely/The most I have ever felt," and valence 0 = "Extremely unpleasant," 100 = "Extremely pleasant."

**Supplementary Methods 4.** Computational modeling

### 1. Computational model

To evaluate gastrointestinal (GI) interoception and participant's behavior in the experimental task, we used a Bayesian model of perception derived from a Markov decision process (MDP) formulation of active inference (Friston, FitzGerald et al. 2017, Friston, Parr et al. 2017, Smith, Friston et al. 2022). Matlab code used to build this model and fit parameters to behavioral data can be accessed at https://github.com/rssmith33/Gut-Inference-Model-Scripts.

Each trial in the model represented a 3-second window during which participants were informed that a vibration could occur. Observations ($o$) were categorical, consisting of vibration, no-vibration, and a formal trial "start" observation. The hidden states ($s$), representing the participant's perception, were also categorical and included vibration, no-vibration, and a "start" state (paired with the "start" observation). Every trial was structured with two timesteps ($t = 1$ and $t = 2$). At $t = 1$, participants always began in the "start" state and made a corresponding "start" observation. At $t = 2$, they either received a vibration or no-vibration observation and inferred whether they had transitioned from the "start" state to either the no-vibration or vibration state. Specifically, by combining their prior beliefs about the transition probabilities from the "start" state $p(s_{t=2}|s_{t=1})$(encoded in a matrix $\mathbf{B}$) with their beliefs about the precision of the mapping between vibration states and observations, $p(o_{t=2}|s_{t=2})$(encoded in a matrix $\mathbf{A}$), they calculated a posterior distribution over states $p(s_{t=2}|o_{t=2})$ as follows:

$$p(s_{t=2}|o_{t=2}) = \boldsymbol{\sigma}\left(\ln \mathbf{B}\, p(s_{t=1}) + \ln \mathbf{A}^{\mathrm{T}}\, o_{t=2}\right) \qquad (4)$$

Here, $\sigma$ indicates a softmax function returning the resulting vector back to a proper probability distribution.

The matrix **B** encodes the probability of transitioning from the "start" state to either the "vibration" ($pV$) or "no vibration" ($1 - pV$) state, as defined in Equation 5. When $pV$ > 0.5, it reflects a prior belief that transitions to the vibration state are more probable (e.g., expecting many vibrations throughout the task). Conversely, when $pV$ < 0.5, it suggests a prior belief that transitions to the "vibration" state are less likely (e.g., expecting a low vibration frequency throughout the task).

$$\mathbf{B} = p(s_{t=2}|s_{t=1}) = \begin{bmatrix} 0 & 0 & 0 \\ 1 - pV & 1 & 0 \\ pV & 0 & 1 \end{bmatrix} \qquad (5)$$

Here, the columns (from left to right) represent the "start" state, "no-vibration" state, and "vibration" state at time $t = 1$, while the rows (from top to bottom) correspond to the same states at time $t = 2$. Additionally, the second and third columns indicate that once an individual reaches either the vibration or no-vibration state, the state remains unchanged within the trial (where a trial is defined by the 3-second time window in which a vibration either was or was not presented).

The matrix **A** encoded the likelihood of receiving a vibration ($IP$) or no-vibration ($1 - IP$) observation, given the presence of the no-vibration or vibration state (Equation 6). When $IP = 0.5$, this indicates minimal precision, meaning that the probability of detecting a vibration or no-vibration is 0.5, regardless of the actual state. Conversely, an $IP$ value approaching 1 reflects high precision, where the probability of perceiving a vibration is high in the vibration state and low in the no-vibration state (and vice versa for no-vibration).

$$\mathbf{A} = p(o_t|s_t) = \begin{bmatrix} 1 & 0 & 0 \\ 0 & IP & 1-IP \\ 0 & 1-IP & IP \end{bmatrix} \tag{6}$$

Similarly, the columns (from left to right) represent the "start" state, "no-vibration" state, and "vibration" state, while the rows (from top to bottom) correspond to the "start" observation, "no-vibration" observation, and "vibration" observation. The likelihood of perceiving either a vibration or no-vibration given the actual state was determined by the interoceptive precision ($IP$) parameter. When $IP$ = 0.5, it indicates minimal precision, meaning that the probability of detecting a vibration or no-vibration is 0.5, regardless of the actual state. Conversely, an $IP$ value approaching 1 reflects high precision, where the probability of perceiving a vibration is high in the vibration state and low in the no-vibration state (and vice versa for no-vibration).

At each trial, the participant response involved two possible actions: pressing or not pressing the button. Our model assumed that the probability of choosing to press the button reflected the posterior probability assigned to the vibration state (versus the no vibration state) at time $t = 2$, as defined in Equation 7:

$$P(press) = \mathbf{P}(\boldsymbol{s_{t=2}} = \boldsymbol{vibration}) \tag{7}$$

In other words, button press behaviors were sampled from the posterior distribution over vibration vs. no vibration states, such that choices to press became more likely as the posterior probability of a vibration approached 1 and choices not to press became more likely as the posterior probability of a vibration approached 0.

Our model also considered the existence of learning processes, meaning that participants could update their prior beliefs about the probability of feeling a vibration vs. no vibration after each trial, based on how frequently they believed they had felt a vibration in the past. Essentially, every time a vibration is felt, prior beliefs favoring feeling a vibration go up, and every time that no vibration is felt this (relative) belief goes back down. Formally, this corresponds to updating the concentration parameters of Dirichlet ($Dir$) priors associated with the **B** matrix ($\boldsymbol{b}$) that specify beliefs about state transitions (Equation 8).

$$p(\boldsymbol{B}) = Dir(\boldsymbol{b})$$

$$pb = \begin{bmatrix} 0 & 0 & 0 \\ 1-pV & 1 & 0 \\ pV & 0 & 1 \end{bmatrix} \tag{8}$$

$$\boldsymbol{b}_{trial} = pb + (1-\omega) \cdot (\boldsymbol{b}_{trial-1} - pb) + \eta \cdot (S_{t=2} \otimes S_{t=1})$$

where ⊗ indicates the cross-product, and $\eta_{pV}$ is a scalar that controls the magnitude of change in concentration parameters after each trial. The parameter $\eta$ acts as a learning rate, controlling the size of the count added to the Dirichlet distribution after each observation. This parameter was estimated separately for vibration and no-vibration trials in the winning model. In some models in our model space, a similar learning process was tested for updating beliefs about $IP$ over time, but this was not found to best account for the data.

### 2. Fitting the model to behavioral data

Our method for parameter estimation utilized a commonly used Bayesian optimization algorithm, known as Variational Laplace (Friston, Mattout et al. 2007), to estimate the computational parameter values for each participant. This algorithm maximized the likelihood of participants' responses, assuming that the posterior probability of perceiving a capsule vibration could be identified with the likelihood of choosing to press the button. We optimized the parameters using this likelihood and variational Laplace (Friston, Mattout et al. 2007), through the *spm_nlsi_Newton.m* parameter estimation routine available in the SPM12 software package (Wellcome Trust Centre for Neuroimaging, London, UK, http://www.fil.ion.ucl.ac.uk/spm). This approach helps mitigate overfitting by imposing complexity cost on parameters that deviate significantly from their prior values.

Estimating parameters required setting prior means and prior variances for each parameter. The prior variance for each parameter was set at a high precision value of 1/4 to help prevent overfitting, and the prior means were assigned as follows (Table S3):

**Table S3.** Prior means for key model parameters

| **Parameter** | **Prior mean value** |
|---|---|
| Interoceptive precision ($IP$) | 0.95 |
| Difference in interoceptive precision ($IP_{diff}$)[§] | 0.25 |
| Initial prior beliefs ($pV$) | 0.5 |
| Learning rates ($\eta_V$ and $\eta_{NV}$) | 0.5 |

[§] Reflects the drop in $IP$ value from the enhanced-intensity to normal-intensity block

Our selection of these priors was informed by our previous study that introduced this approach (Smith, Mayeli et al. 2021). The precision priors assume high precision for the high-intensity block and an intermediate level of precision (.95-.25=0.7) for normal-intensity block.

The prior values for initial prior beliefs ($pV$) and learning rates ($\eta_V$ and $\eta_{NV}$) were specifically selected to reduce estimation bias, with $pV$ = 0.5 reflecting flat initial prior beliefs, and $\eta_V$ and $\eta_{NV}$ equal to 0.5, avoiding any bias toward values near the extremes of 0 or 1.

### 3. Model comparison and computational parameter recoverability

We assessed the relative evidence for multiple models incorporating different parameter combinations (Table S4). In addition to the parameters described in the previous section (Table S3), these models included included variations in learning mechanisms (with different possible learning rates) for updating $pV$. After fitting parameters for each model, we conducted Bayesian model comparison to identify the best-fitting model (Smith, Friston et al. 2022).
Once the optimal model was selected, we verified the recoverability of the associated parameters. Specifically, we simulated behavior using a range of parameter value combinations representative of each participant, estimated parameters from the simulated data, and assessed correlations between generative and estimated parameters. The high correlation between these values confirmed the robustness and recoverability of each parameter.

**Table S4.** Overview of model variants and their corresponding parameters

| $Parameter$ | $IP$ | $IP_{diff}$ | $pV$ | $\eta_{IP}$ | $\eta_{pV}$ |
|---|---|---|---|---|---|
| Model 1 | ✓ | | ✓ | | |
| Model 2 | ✓ | ✓ | ✓ | | |
| Model 3 | ✓ | | ✓ | | ✓ |
| Model 4 | ✓ | ✓ | ✓ | | ✓ |
| Model 5 | ✓ | | ✓ | | ✓\|✓ |
| Model 6* | ✓ | ✓ | ✓ | | ✓\|✓ |
| Model 7 | ✓ | | ✓ | ✓ | |
| Model 8 | ✓ | | ✓ | ✓ | ✓ |
| Model 9 | ✓ | | ✓ | ✓ | ✓\|✓ |
| Model 10 | ✓ | | ✓ | ✓\|✓ | |
| Model 11 | ✓ | | ✓ | ✓\|✓ | ✓ |
| Model 12 | ✓ | | ✓ | ✓\|✓ | ✓\|✓ |

✓ indicates that the corresponding model includes the parameter represented by the respective column; ✓|✓ indicates that the learning rate was split, with separate learning rates for vibration and no-vibration trials; $IP$: interoceptive precision; $IP_{diff}$: difference in IP between normal- and enhanced-intensity stimulations; $pV$: initial prior beliefs; $\eta_{IP}$: learning rate for $IP$ values; $\eta_{pV}$: learning rate for $pV$ values; * indicates the winning model (see results below).

**Supplementary Methods 5.** Processing of peripheral physiological data

1. **Electrogastrogram data**

The single-channel electrogastrographic (EGG) data were processed using custom MATLAB scripts (MATLAB 2021a, MathWorks®) in combination with functions from the FieldTrip toolbox (version 20171022; (Oostenveld, Fries et al. 2011)) and publicly available code developed by Wolpert et al. (2020) (https://github.com/niwolpert/EGG_Scripts/tree/master; (Wolpert, Rebollo et al. 2020)).
The raw EGG signal, originally recorded at 1000 Hz, was downsampled to 250 Hz. It was then bandpass-filtered around the dominant gastric frequency using Wolpert's *compute_filter_EGG()* function, which applies a third-order finite impulse response (FIR) filter with a bandwidth of ±0.015 Hz. Artifact rejection followed criteria defined by Wolpert et al. (2020), whereby cycles exceeding ±3 standard deviations of the cycle length distribution or exhibiting nonmonotonic phase changes were identified and removed (Wolpert, Rebollo et al. 2020). Spectral analysis was conducted using a Fast Fourier Transform (FFT) implemented in the *ft_freqanalysis()* function of the Fireldtrip toolbox to extract key features of gastric motility. To quantify gastric activity, absolute power was calculated for four frequency bands: normogastria (2.5–3.5 cpm), tachygastria (3.75–9.75 cpm), bradygastria (0.5–2.25 cpm), and total power (0.5–11 cpm).

2. **Cardiac data**

The electrocardiogram (ECG) data were processed using custom MATLAB scripts (MATLAB 2021a, MathWorks®) in conjunction with the FieldTrip toolbox (version 20171022; (Oostenveld, Fries et al. 2011)).
The raw ECG signal, initially recorded at 1000 Hz, was downsampled to 250 Hz. Interbeat intervals were extracted from R-peak detection, which was performed using custom scripts incorporating MATLAB's *findpeaks()* function. To quantify heart rate (HR) variability, we computed the standard deviation of normal-to-normal R–R intervals (SDNN), a widely used metric reflecting autonomic regulation of HR. Phasic HR, which captures transient fluctuations in HR in response to mechanical stimulation, was calculated as the difference in HR between the 3-second vibration period and the 3-second pre-stimulation period. Tonic HR, also referred to as resting HR, was estimated by averaging HR over consecutive 60-second windows, representing baseline HR levels in the absence of specific stimulation.

3. **Skin conductance data**

Skin conductance data were processed using custom MATLAB scripts (MATLAB 2021a, MathWorks®) alongside the Ledalab Toolbox (version 3.49; (Bach 2014)).
The raw SCR signal was recorded at 1000 Hz and downsampled to 25 Hz. MATLAB's *detrend()* function was then applied to remove slow signal drifts. To quantify SCR responses to gut stimulation, we calculated the log-transformed maximum phasic activity during the 3-second vibration period, relative to the 3-second pre-stimulus period, applying a 200-millisecond smoothing window and a 0.01 microsiemens (μS) threshold (Benedek & Kaernbach, 2010).

**Supplementary Methods 6.** Illness status definitions.

Illness status was assessed remotely at the 6-month follow-up using an operationalized definition based on our previously proposed standardized criteria (Khalsa, Portnoff et al. 2017). This standardized definition incorporates self-reported symptoms, behaviors, Body Mass Index (BMI), and duration.

1. **Symptoms**

The behavior of eating restriction was assessed using responses on the Restraint subscale of the Eating Disorder Inventory-3 (EDI-3), which evaluates preoccupation with weight, excessive dieting concerns, and fear of gaining weight (Garner, Olmstead et al. 1983). Symptom severity was categorized as follows: a score of ≤ 7 indicated non-significant fear (corresponding to an average response of "Never" or "Rarely"), a score > 7 and ≤ 21 indicated the presence of fear (average response of "Sometimes" or "Often"), and a score > 21 indicated significant fear (average response greater than "Often").

2. **Behaviors**

The behavior of restricting was assessed using the Restraint subscale of the Eating Disorder Examination Questionnaire (EDE-Q) (Fairburn and Beglin 2008). Severity was categorized as follows: a score of ≤ 2 indicated non-significant restriction (occurring 5 or fewer days per month), a score > 2 and ≤ 4 indicated the presence of restriction (occurring more than 5 but no more than 22 days per month), and a score > 4 indicated significant restriction (occurring more than 22 days per month).
Similarly, bingeing and purging behaviors were evaluated using the Bulimia subscale of the Eating Disorder Inventory-3 (EDI-3) (Garner, Olmstead et al. 1983). Scores were interpreted as follows: ≤ 8 indicated non-significant bingeing/purging ("Never" or "Rarely"), > 8 and ≤ 24 indicated the presence of bingeing/purging ("Sometimes" or "Often"), and > 24 indicated significant bingeing/purging (more frequent than "Often").
Purging behaviors involving vomiting or laxative use were further examined using items 26 and 32 from the Body Shape Questionnaire (BSQ) (Cooper, Taylor et al. 1987). A score of 1 ("Never") indicated non-significant purging, a score of 2 or 3 ("Rarely" or "Sometimes") indicated the presence of purging, and a score ≥ 4 ("Often") indicated significant purging.
Finally, excessive exercise as a purging behavior was assessed using item 4 of the Exercise Addiction Inventory (EAI) (Terry, Szabo et al. 2004). Scores were categorized as follows: ≤ 3 indicated no increase in exercise, a score of 4 indicated a present increase, and a score of 5 indicated a significant increase in exercise over time.

3. **Weight and shape concerns**

Weight and shape concerns were evaluated using responses on the Weight Concern and Shape Concern subscales of the EDE-Q. These subscales were selected to avoid overemphasizing or duplicating the Restraint subscale, which was separately operationalized to define restricting behaviors. Specifically, average subscale scores within 1.5 standard deviations (SD) of the HC sample were considered indicative of partial recovery (in combination with the aforementioned symptom and behavior score cutoffs). Scores within 2 SD suggested either full or partial remission, while scores ≥ 2 SD were consistent with partial or full relapse.

4. **Body Mass Index**

BMI was calculated using self-reported height and weight data collected through the EDE-Q. While we asked participants to provide this information, it was not a requirement of the remote follow-up data collection. At the 6-month follow-up, 87% of participants self-reported the data necessary for BMI computation.

Based on these definitions, each AN participant's illness status at 6 months was classified as one of the following:

**Full relapse:** defined as the presence of significant fear of weight gain (EDI-3 Drive for Thinness subscale score > 21), significant restricting behaviors (EDE-Q Restraint subscale score > 4), the presence of bingeing and/or purging behaviors (EDI-3 Bulimia subscale score > 24, BSQ items 26 and 32 scored > 3, and EAI item 4 scored > 4), EDE-Q Weight Concern and Shape Concern subscale averages ≥ 2 SD above those of the HC sample, and a BMI ≤ 18.5.

**Partial relapse:** defined as the presence of significant fear of weight gain (EDI-3 Drive for Thinness subscale score > 21), the presence of restricting behaviors (EDE-Q Restraint subscale score > 2 and ≤ 4), bingeing and/or purging behaviors (EDI-3 Bulimia subscale score > 8 and ≤ 24, BSQ items 26 and 32 scored ≤ 3, and EAI item 4 scored ≤ 4), EDE-Q Weight Concern and Shape Concern subscale averages ≥ 2 SD above those of the HC sample, and a BMI ≤ 18.5.

**Partial remission:** defined using the same criteria as full remission, with the exception that the BMI requirement was ≥ 18.5 but < 19.

**Full remission:** defined as the presence of fear of weight gain (EDI-3 Drive for Thinness subscale score > 7 and ≤ 21), but without restricting behaviors (EDE-Q Restraint subscale score ≤ 2), bingeing or purging behaviors (EDI-3 Bulimia subscale score ≤ 8, BSQ items 26 and 32 scored as 1, and EAI item 4 score ≤ 3). Additionally, individual with AN needed to have EDE-Q Weight Concern and Shape Concern subscale averages within 2 SD of the healthy control (HC) sample and a BMI ≥ 19.

**Partial recovery:** defined as the absence of significant fear of weight gain (EDI-3 Drive for Thinness subscale score ≤ 7), no restricting behaviors (EDE-Q Restraint subscale score ≤ 2), no bingeing or purging behaviors (EDI-3 Bulimia subscale score ≤ 8, BSQ items 26 and 32 scored as 1, and EAI item 4 score ≤ 3), EDE-Q Weight Concern and Shape Concern subscale averages within 1.5 SD of the HC sample, and a BMI ≥ 19.

**Full recovery:** this was not included as a possible 6-month outcome due to the duration requirement of 1 year for this classification.

**Supplementary Methods 7.** Statistical analysis

1. **Behavioral data, computational, and peripheral physiological data**

To assess whether vibratory capsule-induced signals influenced behavioral measures of GI interoception and peripheral physiological responses (cardiac and EGG signals), we employed linear mixed-effects (LME) models. These models evaluated the effects of diagnostic group (AN vs. HC), block (normal vs. enhanced), and period (baseline, normal, enhanced) on the dependent variables. LME models included diagnostic group, block or period, age, and BMI as fixed effects, with participant modeled as a random factor, and measures of GI interoception and peripheral physiological measures as dependent variables. Interactions between diagnostic group and block or period were explicitly tested. To reduce the risk of Type I errors, models were estimated using Restricted Maximum Likelihood (REML), and p-values were derived via Type II analysis of variance (ANOVA) with Satterthwaite's method (Satterthwaite 1941, Luke 2017). Post-hoc analyses were conducted using estimated marginal means, with Kenward-Roger degrees of freedom (Kenward and Roger 1997). The confidence levels and p values were adjusted using a mvt method. Effect size estimates are reported as partial Eta squared ($\eta_p^2$) for Type II ANOVA and as Cohen's d for post-hoc tests.
We also employed linear models to examine whether computational measures were influenced by capsule vibration-induced changes in gastric signals, considering diagnostic group as a key factor. These measures were calculated across both blocks, with age and BMI included as covariates.
Statistical analyses were conducted in R (version 4.2.1; (R Core Team 2024)) using the *lme4* (Bates, Mächler et al. 2015), *lmer* (Kuznetsova, Brockhoff et al. 2017), and *emmeans* packages.

2. **Electroencephalographic data**

We applied non-parametric cluster permutation testing to examine whether gut mechanosensory stimulations induced by capsule vibration influenced vibration-elicited Event-Related Potential (ERP) amplitudes as a function of diagnostic group (AN vs. HC). This data-driven statistical method allowed for group comparisons across all electrodes and time points, offering a robust approach to handling the multiple comparison problem (Maris and Oostenveld 2007, Maris 2012). The underlying assumption of this approach is that a biologically meaningful effect should exhibit spatial and temporal coherence across neighboring electrodes and time points. Statistical significance was determined by comparing the observed statistical results with a distribution of values generated through permutation-based shuffling of the data (Maris and Oostenveld 2007, Maris 2012). Given the absence of prior assumptions regarding the spatial and temporal distribution of ERP differences between diagnostic groups, this exploratory approach was particularly well suited to our study.
Cluster identification involved the following steps: (a) t-statistics were calculated for each sample in the spatiotemporal ERP data to compare diagnostic groups (AN vs. HC); (b) these statistics were thresholded by a p-value ($p < 0.05$); (c) neighboring data points that exceeded the threshold and had the same sign were grouped; (d) cluster-level statistics were obtained by selecting the maximum t-statistic within each cluster; and (e) the maximum cluster statistic was assessed against its permutation distribution. This permutation distribution was derived from statistical values of independent t-tests performed on 10,000 random permutations of ERP data relative to diagnostic group, with a p-value threshold of 0.05 for cluster inclusion (Pernet, Latinus et al. 2015). The analysis accounted for both spatial (electrodes) and temporal (time points) dimensions. Electrodes within 2.5 cm were considered neighbors, averaging 5.4 neighbors per electrode, and a minimum of two neighboring electrodes was required for a sample to be included in the clustering process.

The analysis was implemented using custom scripts in Matlab 2021a and the Fieldtrip toolbox (version 20171022; (Oostenveld, Fries et al. 2011)).

**Supplementary Results 1.** Electroencephalogram findings

### 1. Group comparisons of onset-evoked ERP responses

Cluster-based permutation testing did not reveal significant group differences in vibration onset-evoked ERP amplitude across blocks (normal block: Monte Carlo P>0.27; enhanced block: Monte Carlo P>0.13; Figure S2).

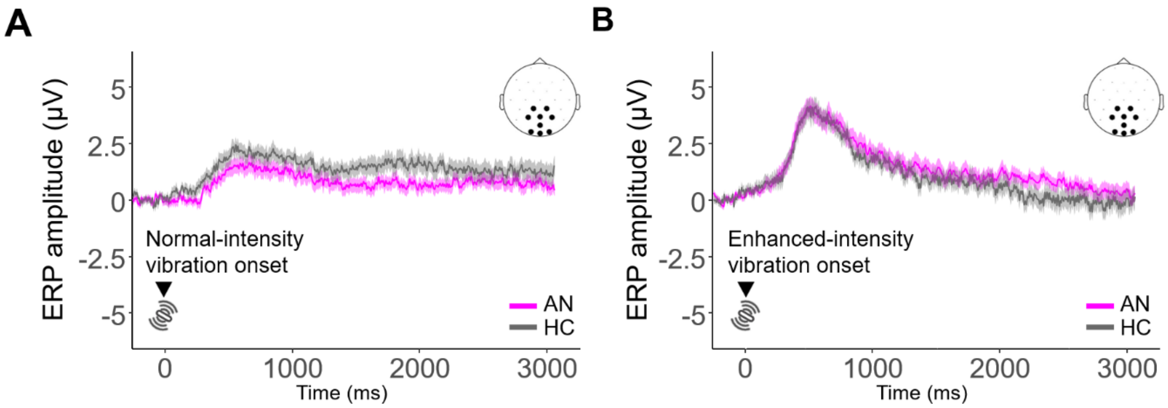


**Figure S2 – Group overlay of onset-evoked event-related potential (ERP) responses.** There were no significant differences in ERP amplitude between the AN and HC groups during the normal block **(A)** and the enhanced block **(B)**. Error bars indicate standard error of the mean. *Note: The ERP was computed from the cluster of centro-right parieto-occipital electrodes exhibiting a significant intensity-vibration (block) effect in both groups.* AN – anorexia nervosa, HC – healthy comparison.

### 2. Group comparisons of offset-evoked ERP responses

Vibration offset-locked analyses revealed similar results. We did not find significant group differences in ERP amplitude across blocks (normal block: Monte Carlo P>0.20; enhanced block: Monte Carlo P>0.34; Figure S3).

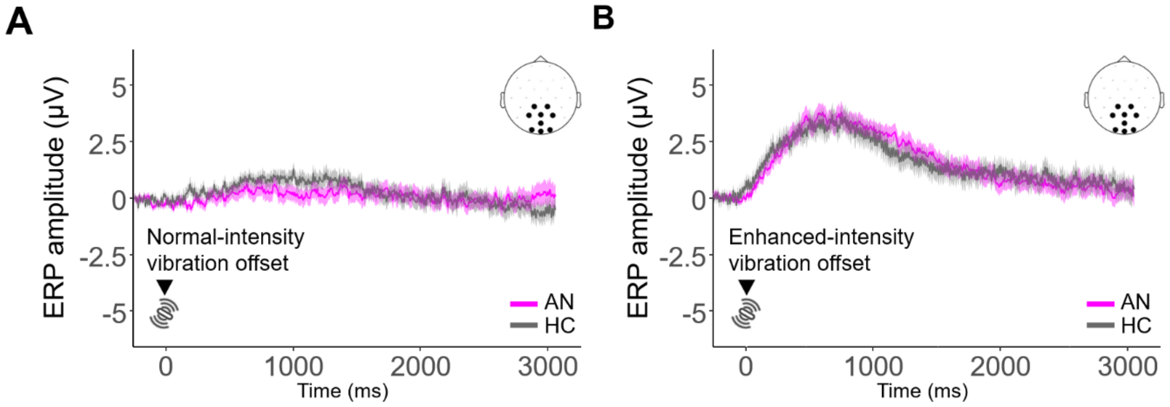


**Figure S3 – Group overlay of offset-evoked event-related potential (ERP) responses.** ERP amplitudes did not differ significantly between the AN and HC groups during the normal block **(A)** or the enhanced block **(B)**. Error bars indicate standard error of the mean. *Note: The ERP was computed from the cluster of centro-right parieto-occipital electrodes exhibiting a significant intensity-vibration (block) effect in both groups.* AN – anorexia nervosa, HC – healthy comparison.

**Supplementary Results 2.** Model comparison and parameter recoverability

Table S5 presents the results of the model comparison. A clear winning model emerged (Model 6 with a protected exceedance probability of 1), which included the following parameters: $IP$, $IP_{diff}$, $pV$, and separate learning rates for priors ($\eta_{pV}$) when observing versus not observing a vibration (referred to as $\eta_V$ vs. $\eta_{NV}$).

**Table S5.** Model comparison results

| | **Protected exceedance probability** |
|---|---|
| Model 1 | $4.09\times10^{-43}$ |
| Model 2 | $4.09\times10^{-43}$ |
| Model 3 | $4.09\times10^{-43}$ |
| Model 4 | $4.09\times10^{-43}$ |
| Model 5 | $4.09\times10^{-43}$ |
| Model 6 | 1 |
| Model 7 | $4.09\times10^{-43}$ |
| Model 8 | $4.09\times10^{-43}$ |
| Model 9 | $4.09\times10^{-43}$ |
| Model 10 | $4.09\times10^{-43}$ |
| Model 11 | $4.09\times10^{-43}$ |
| Model 12 | $4.09\times10^{-43}$ |

Recoverability analyses confirmed that these parameters were reliably recoverable within the range of values observed in participant estimates. Specifically, when generating simulated behavior using the parameter value combinations observed in participant estimates and then re-estimating the parameters from the simulated data, the correlations between true and estimated parameters were strongly positive (Table S6).

**Table S6.** Correlation results for parameter recoverability analysis

| Parameter | Correlation between true and estimated parameters | |
|---|---|---|
| | *r* | P-value |
| Interoceptive precision ($IP$) | 0.91 | <0.001 |
| Difference in interoceptive precision ($IP_{diff}$) | 0.97 | <0.001 |
| Initial prior beliefs ($pV$) | 0.64 | <0.001 |
| Learning rate for trials with capsule vibration ($\eta_V$) | 0.77 | <0.001 |
| Learning rate for trials without capsule vibration ($\eta_{NV}$) | 0.92 | <0.001 |

**Supplementary Results 3.** Peripheral physiological findings

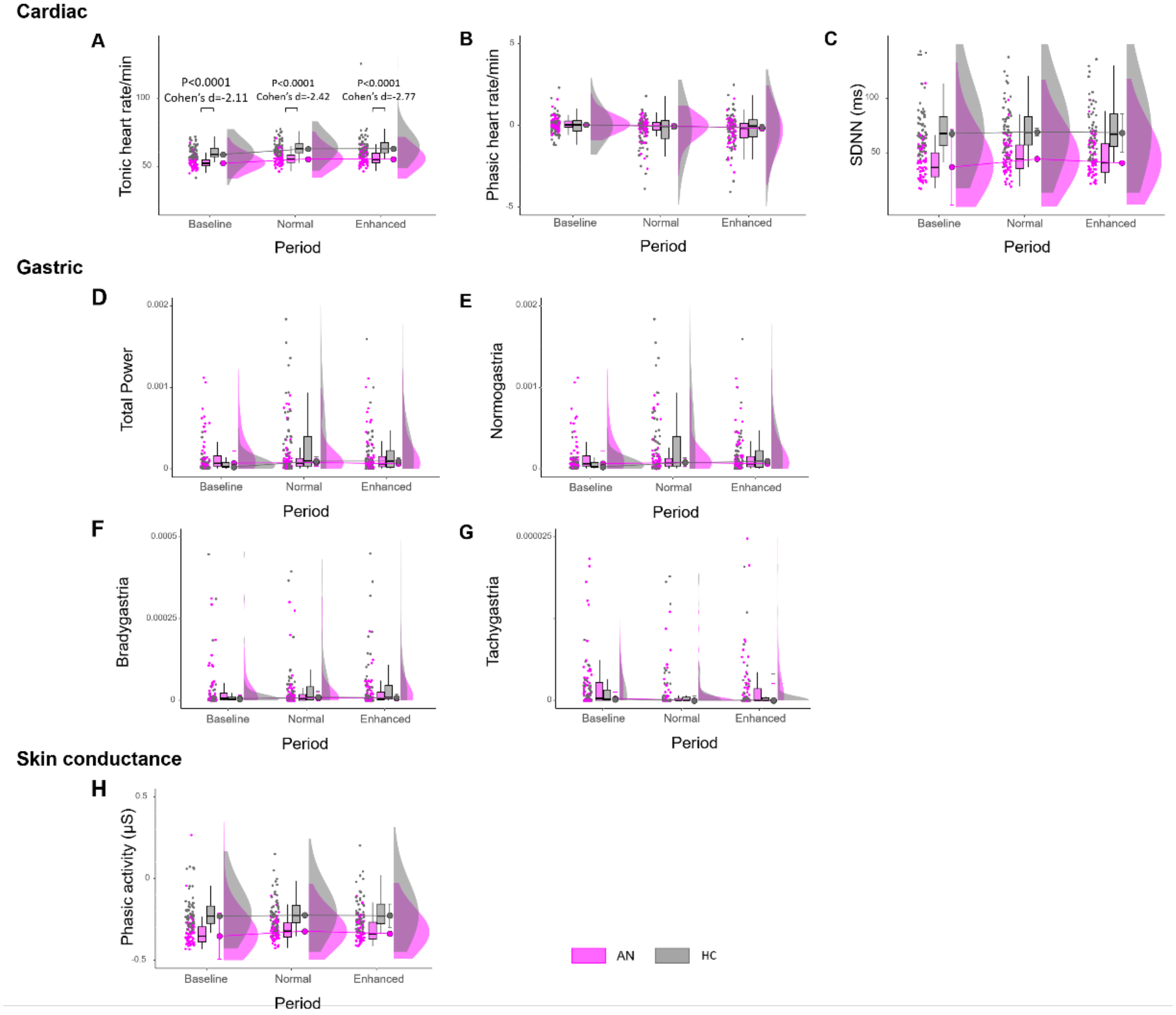


**Figure S4 – Peripheral physiological responses during baseline and vibratory gut stimulation. (A)** The AN group exhibited a significantly lower tonic (basal) heart rate (HR) compared to HCs, with group differences becoming more pronounced during the stimulation periods relative to baseline. **(B-C)** No significant differences were observed between groups in phasic HR activity or heart rate variability, assessed via the standard deviation of R–R intervals (SDNN). **(D-E-F-G)**. Similarly, no group differences were found in electrogastrogram (EGG) responses, including absolute power across four gastric frequency bands: total power ([0.5–11] cpm), bradygastria ([0.5–2.25] cpm), normogastria ([2.5–3.5] cpm), and tachygastria ([3.75–9.75] cpm). **(H)** Skin conductance response (SCR) also showed no significant differences between groups, particularly in the maximum value of phasic activity. The plots display the distribution of individual data points, corresponding boxplots, and individual participant values. AN – Anorexia Nervosa, HC – healthy comparison, ms – milliseconds, μS – microsiemens. *Note: Only significant main group effects are reported in the figures.*

**Supplementary Results 4. Longitudinal findings**

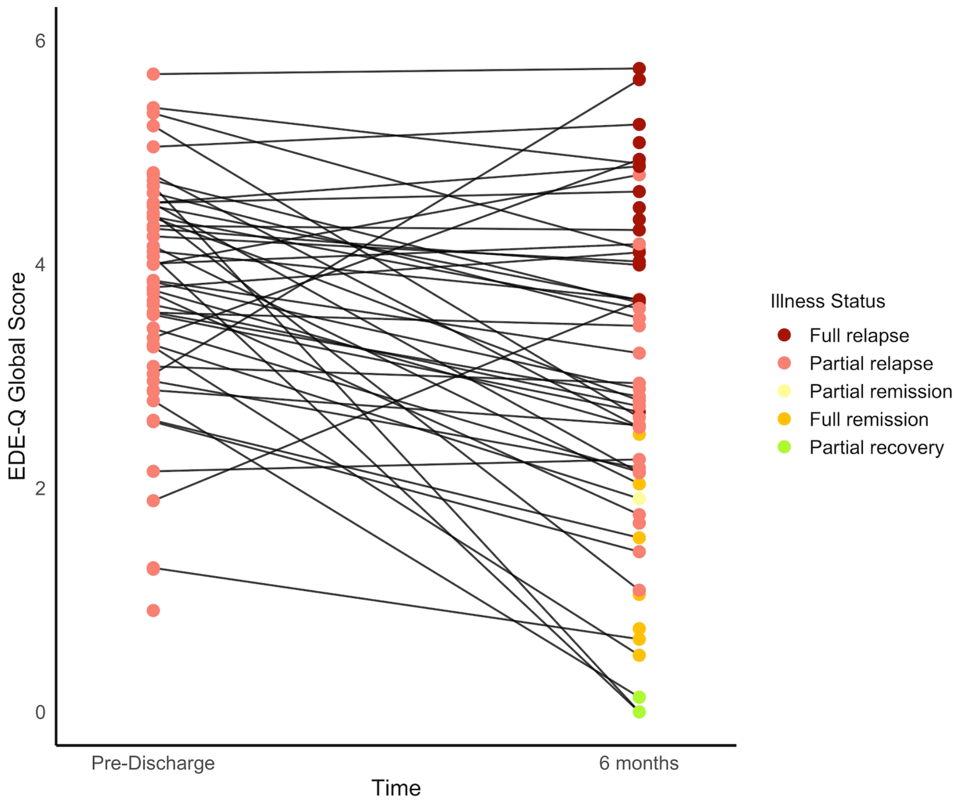


**Figure S5.** Spaghetti plot of AN participants showing eating disorder symptoms (indexed via the EDE-Q Global Score) at the baseline pre-discharge visit and again at the 6-month follow-up visit. Lower EDE-Q Global Scores indicate lower eating disorder symptomatology. The color coding of each individual's clinical status at each visit timepoint corresponds to the operationalized definitions for relapse, remission and partial recovery adapted from our previously proposed standardized criteria (Khalsa, Portnoff et al. 2017).

**Table S7.** Logistic regression results predicting 6-month AN status (full relapse) from experimental session measures. Statistically significant predictions are bolded, with the corresponding Odd Ratio (OR) values reported.

| **Predictor** | **Outcome 6-month AN status** | |
|---|---|---|
| | P-value | OR |
| Interoceptive precision ($IP$) | 0.20 | - |
| Difference in interoceptive precision ($IP_{diff}$) | 0.10 | - |
| Initial prior beliefs ($pV$) | **0.05** | **3.82** |
| Learning rate for trials with capsule vibration ($\eta_V$) | 0.71 | - |
| Learning rate for trials without capsule vibration ($\eta_{NV}$) | 0.45 | - |
| Normalized A' (Normal block) | 0.77 | - |
| Normalized A' (Enhanced block) | 0.74 | - |
| Miss rate (Normal block) | 0.64 | - |
| Miss rate (Normal block) | 0.36 | - |
| Response bias (Normal block) | **0.04** | **5.37** |
| Response bias (Enhanced block) | 0.94 | - |
| Response time (Normal block) | 0.18 | - |
| Response time (Enhanced block) | 0.82 | - |

**Table S8.** Linear regression results predicting EDE-Q Total score at 6 months from experimental session measures. Statistically significant predictions are bolded, with the corresponding Adjusted R squared ($R^2$) values reported.

| **Predictor** | **Outcome 6-month EDE-Q Total score** | |
|---|---|---|
| | P-value | $R^2$ |
| Interoceptive precision ($IP$) | 0.61 | - |
| Difference in interoceptive precision ($IP_{diff}$) | **0.004** | **0.16** |
| Initial prior beliefs ($pV$) | **0.05** | **0.09** |
| Learning rate for trials with capsule vibration ($\eta_V$) | 0.06 | - |
| Learning rate for trials without capsule vibration ($\eta_{NV}$) | 0.25 | - |
| Normalized A' (Normal block) | 0.23 | - |
| Normalized A' (Enhanced block) | 0.29 | - |
| Miss rate (Normal block) | **0.05** | **0.08** |
| Miss rate (Enhanced block) | 0.46 | - |
| Response bias (Normal block) | 0.14 | - |
| Response bias (Enhanced block) | 0.80 | - |
| Response time (Normal block) | 0.53 | - |
| Response time (Enhanced block) | 0.47 | - |

**Supplementary Results 5.** Multilevel correlation findings

**Table S9.** Correlations between average ERP (avERP) amplitude and behavioral or computational measures for the normal and enhanced stimulation blocks. Statistically significant correlations (p<0.05), following Holm correction, are bolded. *Note: The avERP amplitude was derived from the spatiotemporal window identified as the electroencephalographic marker of gut mechanosensation across diagnostic groups (individuals with anorexia nervosa (AN; n=62) and healthy comparisons (HCs; n=57); see Figures 3A-B in the main text).*

| | **Correlation with the avERP amplitude** | | | |
|---|---|---|---|---|
| | **Normal** | | **Enhanced** | |
| **Measure** | **AN** | **HC** | **AN** | **HC** |
| Normalized A' | ***r*=0.70**<br>**p<0.001** | ***r*=0.49**<br>**p=0.04** | *r*=0.20<br>p=1 | *r*=0.33<br>p=1 |
| Miss rate | ***r*=-0.66**<br>**p<0.001** | ***r*=-0.57**<br>**p<0.001** | *r*=-0.21<br>p=1 | *r*=-0.34<br>p=1 |
| Response time | ***r*=-0.60**<br>**p=0.002** | ***r*=-0.68**<br>**p<0.001** | *r*=-0.08<br>p=1 | *r*=-0.34<br>p=1 |
| Response bias | *r*=-0.01<br>p=1 | *r*=-0.16<br>p=1 | *r*=-0.15<br>p=1 | *r*=-0.27<br>p=1 |
| Initial prior beliefs ($pV$) | *r*=0.26<br>p=1 | *r*=0.27<br>p=1 | *r*=0.07<br>p=1 | *r*=0.01<br>p=1 |
| Interoceptive precision ($IP$) | *r*=0.27<br>p=0.11 | *r*=0.11<br>p=1 | *r*=0.21<br>p=1 | *r*=0.36<br>p=1 |
| Difference in interoceptive precision ($IP_{diff}$) | ***r*=-0.62**<br>**p<0.001** | *r*=-0.37<br>p=0.69 | *r*=-0.19<br>p=1 | *r*=-0.14<br>p=1 |
| Learning rate for trials with capsule vibration ($\eta_V$) | ***r*=0.58**<br>**p<0.001** | ***r*=0.49**<br>**p=0.01** | *r*=0.37<br>p=0.36 | *r*=0.22<br>p=1 |
| Learning rate for trials without capsule vibration ($\eta_{NV}$) | ***r*=-0.60**<br>**p<0.01** | ***r*=-0.46**<br>**p=0.04** | *r*=-0.38<br>p=0.34 | *r*=-0.25<br>p=1 |

**Supplementary Results 6.** Self-report findings

**Table S10.** Self-report measures: logistic regression predicting 6-month AN illness status (full relapse) from retrospective ratings of sensation intensity and valence (unpleasant/pleasant) and state items before, during, and after vibratory gut stimulation. Statistically significant predictions are bolded, with the corresponding Odds Ratio (OR) values reported.

| **Predictor** | **Outcome 6-month full relapse status** | |
|---|---|---|
| | P-value | OR |
| Intensity of sensations | *Before* \| *During* | *Before* \| *During* |
| Stomach/digestive | 0.10 \| 0.55 | - \| - |
| Breath | 0.11 \| 0.48 | - \| - |
| Heartbeat | 0.21 \| 0.49 | - \| - |
| Muscle tension | 0.69 \| 0.81 | - \| - |
| Valence of sensations | *Before* \| *During* | *Before* \| *During* |
| Stomach/digestive | 0.38 \| **0.03** | - \| **5.73** |
| Breath | 0.87 \| 0.67 | - \| - |
| Heartbeat | 0.48 \| 0.43 | - \| - |
| State | *Before* \| *After* | *Before* \| *After* |
| Hunger | 0.19 \| 0.49 | - \| - |
| Thirst | 0.21 \| 0.22 | - \| - |
| Urge to urinate | 0.31 \| 0.90 | - \| - |
| Urge to defecate | 0.10 \| 0.62 | - \| - |

**Table S11.** Self-report measures: linear regression results predicting EDE-Q Total score at 6 months from retrospective ratings of sensation intensity and valence (unpleasant/pleasant) and state items before, during, and after vibratory gut stimulation. Statistically significant predictions are bolded, with the corresponding Adjusted R squared ($R^2$) values reported.

| **Predictor** | **Outcome 6-month EDE-Q Total score** | |
|---|---|---|
| | P-value | $R^2$ |
| Intensity of sensations | *Before* \| *During* | *Before* \| *During* |
| Stomach/digestive | **0.01** \| 0.83 | **0.13** \| - |
| Breath | 0.17 \| 0.48 | - \| - |
| Heartbeat | 0.16 \| 0.97 | - \| - |
| Muscle tension | 0.81 \| 0.09 | - \| - |
| Valence of sensations | *Before* \| *During* | *Before* \| *During* |
| Stomach/digestive | **0.02** \| **0.02** | **0.11** \| **0.11** |
| Breath | 0.67 \| 0.73 | - \| - |
| Heartbeat | 0.97 \| 0.34 | - \| - |
| State | *Before* \| *After* | *Before* \| *After* |
| Hunger | 0.73 \| 0.62 | - \| - |
| Thirst | 0.50 \| 0.45 | - \| - |
| Urge to urinate | 0.71 \| 0.97 | - \| - |
| Urge to defecate | 0.28 \| 0.48 | - \| - |
